# Longitudinal changes of white matter hyperintensities in sporadic small vessel disease: a systematic review and meta-analysis

**DOI:** 10.1101/2022.03.22.22272696

**Authors:** Angela CC Jochems, Carmen Arteaga, Francesca M Chappell, Tuula Ritakari, Monique Hooley, Fergus N Doubal, Susana Muñoz Maniega, Joanna M Wardlaw

**Affiliations:** Centre for Clinical Brain Sciences and UK Dementia Research Institute at the University of Edinburgh, Edinburgh, United Kingdom; Centre for Discovery Brain Sciences, University of Edinburgh, Edinburgh, United Kingdom

**Author notes:** **Corresponding author:** Joanna M Wardlaw.

## Abstract

**Background and objectives:** White matter hyperintensities (WMH) are frequent imaging features of small vessel disease (SVD) and related to poor clinical outcomes. WMH progression over time is well described, but regression was also noted recently, although the frequency and associated factors are unknown. This systematic review and meta-analysis aims to assess longitudinal intra-individual WMH volume changes in sporadic SVD.

**Methods:** Following PRISMA guidelines we searched EMBASE and MEDLINE for papers up to 28 January 2022 on WMH volume changes using MRI on _≥_2 time-points in adults with sporadic SVD. We classified populations (healthy/community-dwelling, stroke, cognitive, other vascular risk factors, depression) based on study characteristics. We performed random-effects meta-analyses with Knapp-Hartung adjustment to determine mean WMH volume change (change in mL, % of intracranial volume [%ICV], or mL/year), 95%CI and prediction intervals (PI, limits of increase and decrease) using unadjusted data. Risk of Bias Assessment tool for Non-randomised Studies (RoBANS) was used to assess risk of bias.

**Results:** Forty papers, 10,932 participants, met the inclusion criteria. Mean WMH volume increased over time by: 1.74 mL (95% CI 1.23, 2.26; PI -1.24, 4.73 mL; 27 papers, N=7411, mean time interval 2.7 years, SD=1.65); 0.25%ICV (95% CI 0.14, 0.36; PI -0.06, 0.56; 6 papers, N=1071, mean time interval 3.5 years, SD =1.54); or 0.61 mL/year (95% CI 0.37, 0.85; PI -0.25, 1.46; 7 papers, N=2450). Additionally, 13 papers specifically mentioned and/or provided data on WMH regression, which occurred in asymptomatic, stroke, and cognitive presentations of SVD.

**Discussion:** Net mean WMH volume increases over time mask wide-ranging change (e.g. mean increase of 1¾mL ranging from 1¼mL decrease to 4¾ml increase), with regression documented explicitly in up to 1/3 of participants. More knowledge on underlying mechanisms, associated factors and clinical correlates is needed, as WMH regression could be an important intervention target.

## Introduction

White matter hyperintensities (WMH) of presumed vascular origin are the most common neuroimaging feature of small vessel disease (SVD), a disorder of the cerebral microvessels. WMH are visible on MRI as hyperintense lesions on T2-weighted and hypointense on T1-weighted sequences.^1^

Around 11-15% of a general middle-aged population have one or more WMH, increasing to over 90% in people aged over 80. WMH are associated with risk factors including age, hypertension, smoking, and diabetes^2^ and with symptoms such as apathy, fatigue, delirium, cognitive decline and increased risk of stroke and dementia.^3^

WMH are thought to indicate areas of permanent white matter damage due to demyelination and axon loss.^3^ In longitudinal studies, WMH progression over time is a common finding, while WMH regression has only been noted in a few recent studies,^4, 5^ but otherwise has been disregarded as measurement error or overlooked. If a genuine finding, then WMH regression might suggest that WMH do not only indicate permanently damaged brain tissue. A better understanding of the frequency and factors associated with WMH regression could help identify potential interventions to delay WMH progression and the devastating clinical consequences.

We performed this systematic review and meta-analysis to assess longitudinal intra-individual WMH volume changes in sporadic SVD over time using volumetric MRI measurements and progression rates.

## Methods

### Search strategy and selection criteria

We searched EMBASE and MEDLINE from January 1985, when MRI became more widely implemented in clinical practice, to January 28, 2022, for studies investigating longitudinal quantification of WMH volume on MRI on at least two different times points in adults over 18 years old (eAppendix 1: Search strategy). We designed the search strategy and complied with the Preferred Reporting in Systematic Review and Meta-Analysis (PRISMA) guidelines. We supplemented this search with hand-searched articles from January 2012 to December 22, 2020, in Stroke, Journal of Cerebral Blood Flow and Metabolism, and Neurology. We registered the protocol on PROSPERO (CRD42018080548) on January 23, 2018. We included published full-text articles from peer-reviewed longitudinal studies that used MRI to quantify WMH volumes, defined according to STRIVE criteria^1^, on at least two different time points and that provided numerical analysis of WMH volume change between the time points. Studies included randomised trials, non-randomised trials, cohort studies, and case-control studies. For data from a single cohort published more than once, we included the most relevant paper with the largest sample size and years of follow-up and most useable data to minimise duplication or overlapping samples. We excluded studies of SVD attributable to hereditary causes (e.g., CADASIL, CARASIL) or WMH attributable to other causes (e.g., multiple sclerosis, inflammatory disorders including primary angiitis, secondary vasculitis, post-infectious, and paraneoplastic syndromes).

Title and abstract screening and duplicate paper removal were done independently by one reviewer (CA, ACCJ, TR, MH). A second reviewer screened a random 10% sample of titles and abstracts. Full-text review was assessed independently by one reviewer, and a second reviewer screened a random 20% sample of the full texts (CA, ACCJ) using Covidence software (https://www.covidence.org/). Data extraction was performed by a single reviewer using a pre-specified data collection form (eTable 1), and a second reviewer double-extracted a random 20% sample. Two reviewers used RoBANS (risk of bias assessment tool for non-randomised studies)^6^ to assess all studies including randomised trials, for participant selection, adjustment for confounders, adequacy of WMH descriptors, blinding, incomplete outcome data, and selective outcome reporting (eTable 2). Any disagreements were resolved by discussion between reviewers (CA, ACCJ) with the help of a senior reviewer (JMW).

### Data extraction

We extracted information on study design, demographic characteristics, vascular risk factors, study population (i.e. healthy and/or community-dwelling population (healthy/CD), patients with depression, patients with stroke, cognitive presentation or other vascular risk factors (VRF) presentations), and follow-up period.

We extracted data related to MRI assessment, including WMH volumes per time point and WMH change between those time points, adjustment of WMH measurements to e.g., intracranial volume, total brain volume; methods of WMH calculation and predictors of change. WMH volume data are often skewed and are log transformed to normalise the data before being used in analyses. We aimed to use unadjusted raw data where possible to reflect real intra-individual changes and include the whole range of least and most growth. We selected studies that provided unadjusted mean raw WMH volume change, e.g. mL, cm^3^ and cc, changes in volume expressed as a percentage of intracranial volume (%ICV), or an annual change rate (e.g. mL per year). We extracted the mean or median with corresponding SD, IQR, range, 95% CI, SEM, and if provided, the baseline WMH volumes. If studies did not provide mean and SD of WMH volume change, we calculated the mean and standard deviation from the sample size, median, range, IQR, SEM or 95% CI.^7^ We extracted data from studies that reported volumes per group and for the entire cohort; when the volume for the entire cohort was not available, data for the separate groups were included. For studies that did not report complete volumetric assessments, or where the data were not useable in the meta-analysis, we extracted relevant measures to perform a narrative summary of findings.

### Statistical analysis

We used random-effects meta-analyses (because of expected between-study heterogeneity) to calculate mean WMH change, its confidence interval and prediction interval (PI) using untransformed raw means. More than two studies are needed to calculate a PI. The PI is an estimate of an interval, based on data that have already been observed and are included in the meta-analysis, in which future observations will fall, with 95% confidence. In contrast to the CI, it indicates the range of least and most WMH volume change in a sample, while the CI indicates the range of mean change. We applied the Knapp-Hartung adjustment to control for uncertainty regarding between-study heterogeneity and calculate the confidence interval around the pooled mean.^8^ We planned additional explorative subgroup analyses by study population group (e.g. stroke), age strata, and time lapse between MRIs, when possible. Populations were assigned based on the study characteristics. When papers reported subgroups, e.g. cases, controls, or treatment groups, these were entered as separate groups and mentioned in the main meta-analyses unless data from the total group was used. However, there were insufficient data to assess trial interventions. We assessed heterogeneity by visual assessment of the forest plots and by calculating the I² and τ^2^ statistics to estimate the between-study variance with a restricted maximum-likelihood estimator. We used R version 4.0.2 (https://www.R-project.org/) and the ‘meta’ package.^9^

## Data availability

Data used in this study are available upon reasonable request from the corresponding author.

## Results

Our search yielded 1206 publications and our manual search provided 197 further publications (Figure 1). After title and abstract screening, 248 full texts were assessed for eligibility. Most papers were excluded because they only reported WMH volumes at one time point, reported no WMH volumes at all, or were superceded by a publication from the same study reporting more complete relevant data. This left 64 papers that were relevant to the review question, but 24 papers did not provide raw/unadjusted WMH volumes and instead used log-transformed, estimated volumes, or percentages of change compared to baseline volumes. Therefore, 40 papers were included in meta-analysis, comprising 39 different studies and 10,932 participants (summarised in eTables 3-5), time between scans median 2.6 years (range 0.25 – 8.7 years). For 13/40 papers included in the meta-analysis we had to estimate the mean and SD. The 24 relevant papers that did not provide useable WMH change data are summarised in text, eResults 1-2 and eTable 6.

**Figure 1.**
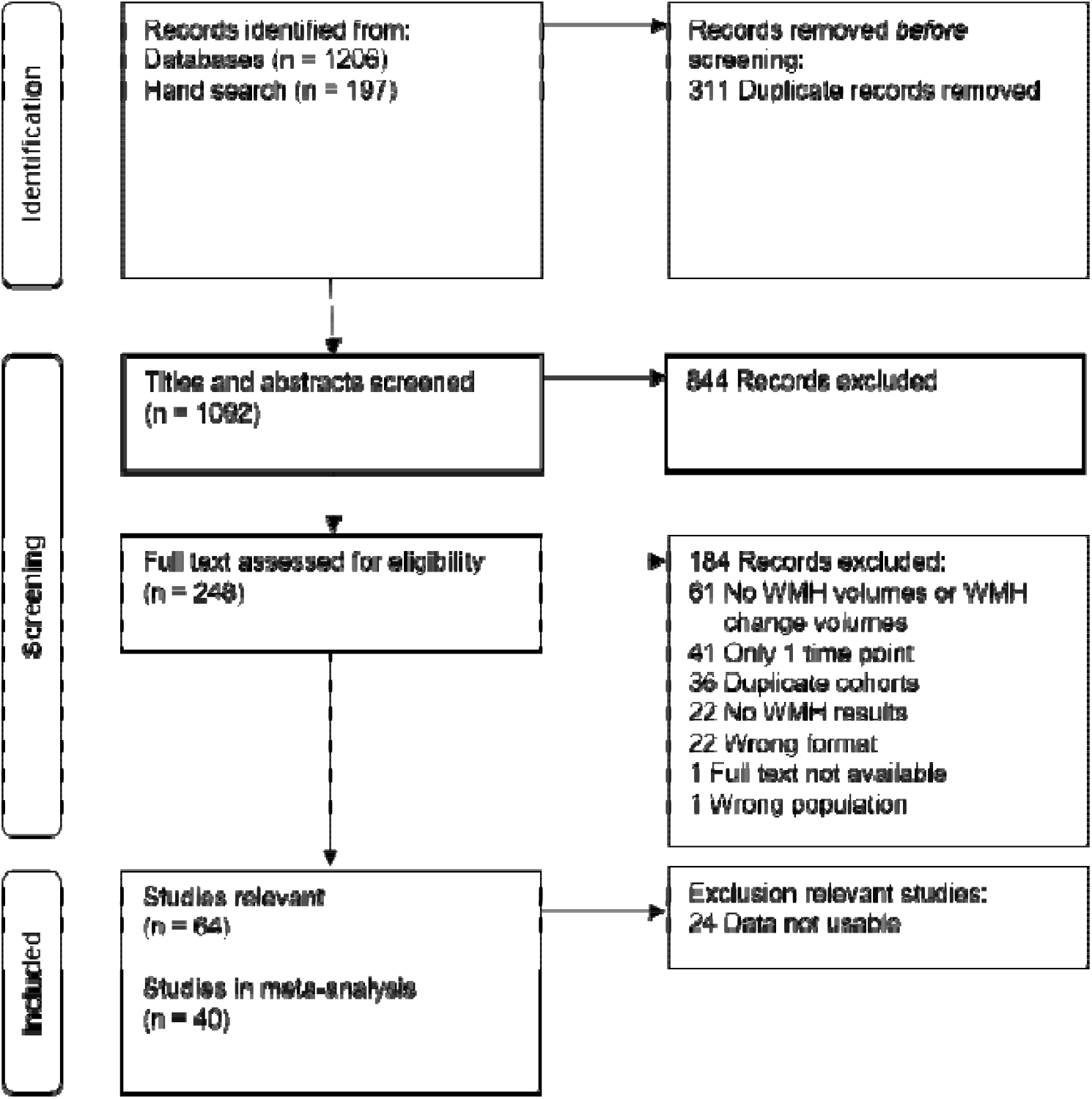
Flow chart of study selection

### Intra-individual WMH change

#### WMH volume change in mL

We identified 27 papers^4, 5, 10-34^ (total 7411 participants) that reported raw WMH volume change over time. Overall time between scans was on average 2.7 years (SD=1.65; median=2 years; range 0.25 - 8.1 years). We combined all data from all populations in one meta-analysis (Figure 2). Overall WMH increased by mean 1.74 mL over time (95% CI 1.23, 2.26 mL), with PI of -1.24, 4.73 mL).

**Figure 2.**
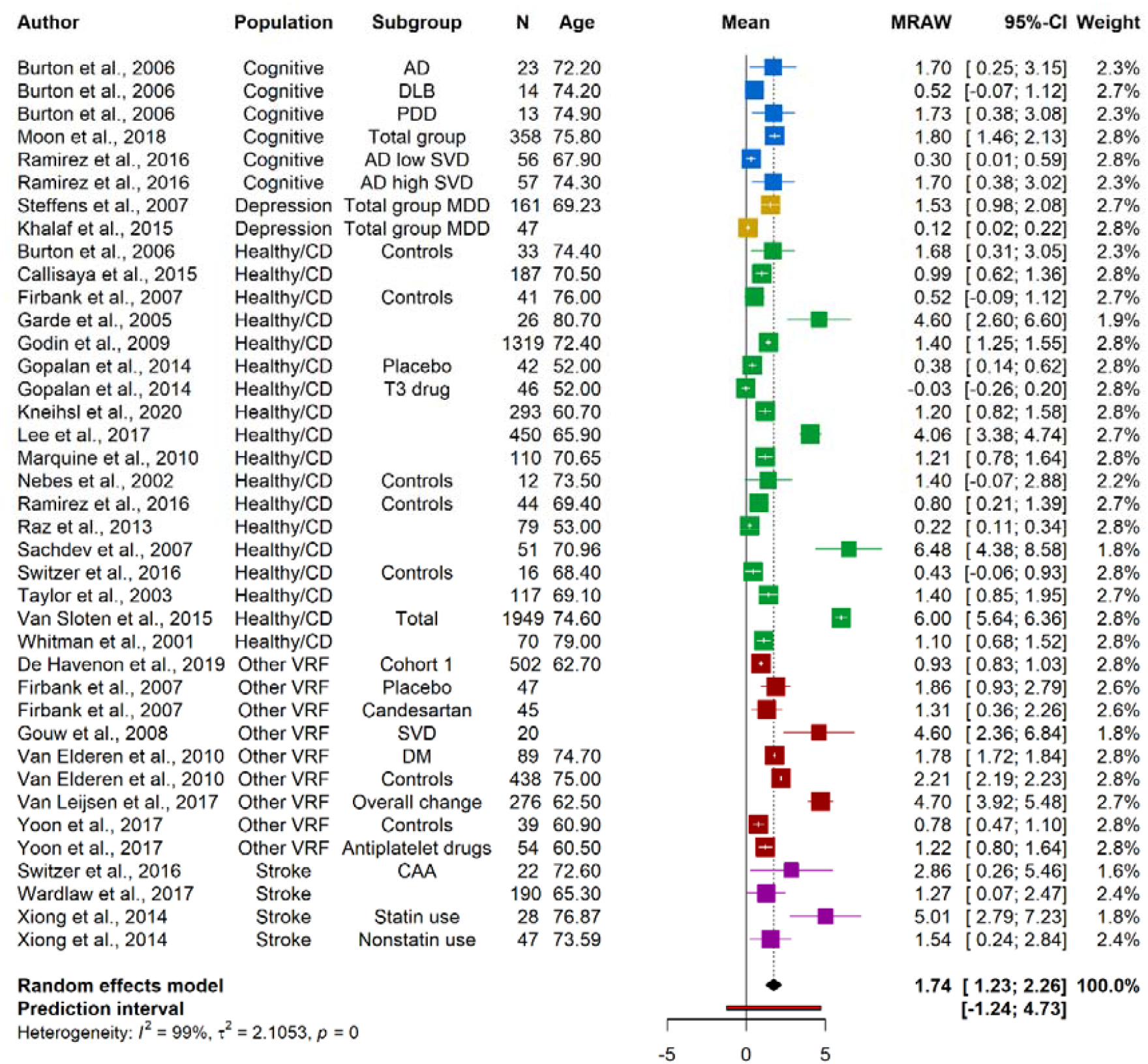
Random-effects meta-analysis of raw mean WMH volume change in mL over median 2 years, range 0.25 - 8.1 years. Squares represent means and bars the 95%CI. Note: AD: Alzheimer’s disease; CAA: Cerebral amyloid angiopathy; DLB: Lewy Body Dementia; DM: Diabetes Mellitus; Healthy/CD: Healthy/community-dwelling; MDD: Major depressive disorder; MRAW: Raw means; PDD: Parkinson’s disease dementia; SVD: Small vessel disease; VRF: Vascular risk factors

#### Change in %ICV

Six studies reported WMH and their change over time as %ICV^35-40^ (total 1071 participants), with time between MRI mean 3.5 years (SD=1.54. Median=3; range 1.9 – 6.7) were meta-analysed (Figure 3). WMH volume as %ICV shows an increase of 0.25 (95% CI 0.14, 0.36;PI -0.06, 0.56) %ICV.

**Figure 3.**
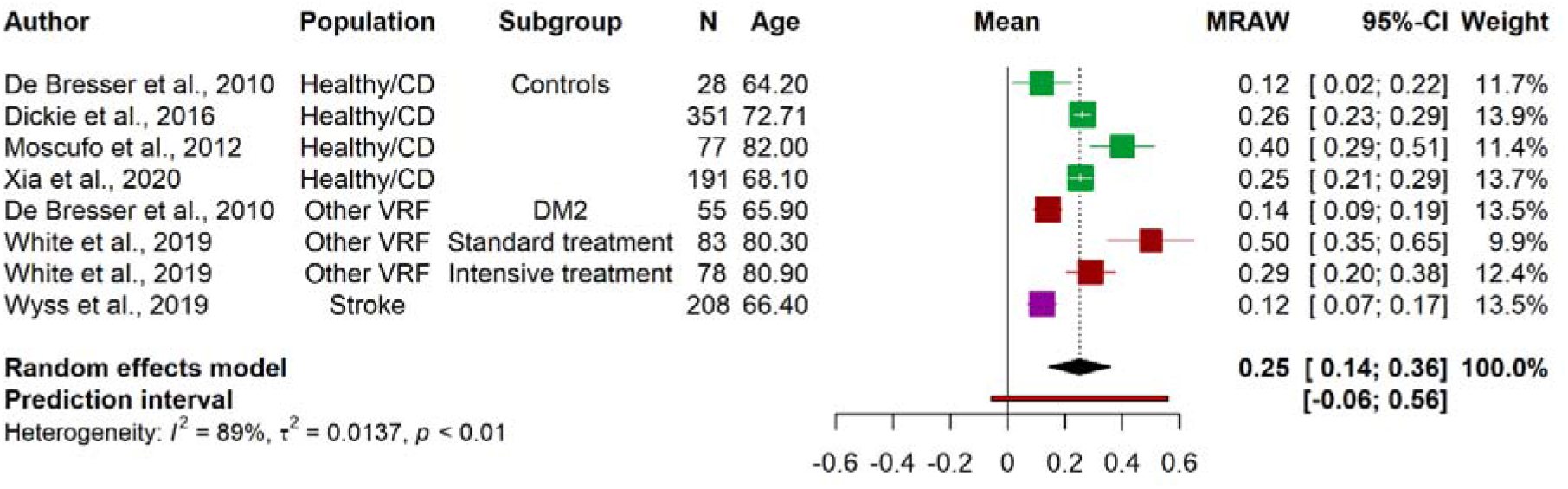
Random-effects meta-analysis of mean WMH volume change as %ICV over median of 3 years, range 1.9 – 6.7 years. Squares represent means and bars the 95%CI. Note: DM2: Diabetes Mellitus type 2; Healthy/CD: Healthy/community-dwelling; MRAW: Raw means; VRF: Vascular risk factors

#### WMH change in mL per year

Seven studies^41-47^ (total 2450 participants) reported the unadjusted mean WMH change per year (Figure 4). Overall, mean WMH change showed an increase of 0.61 (95% CI 0.37, 0.85; PI -0.25, 1.46) ml/year.

**Figure 4.**
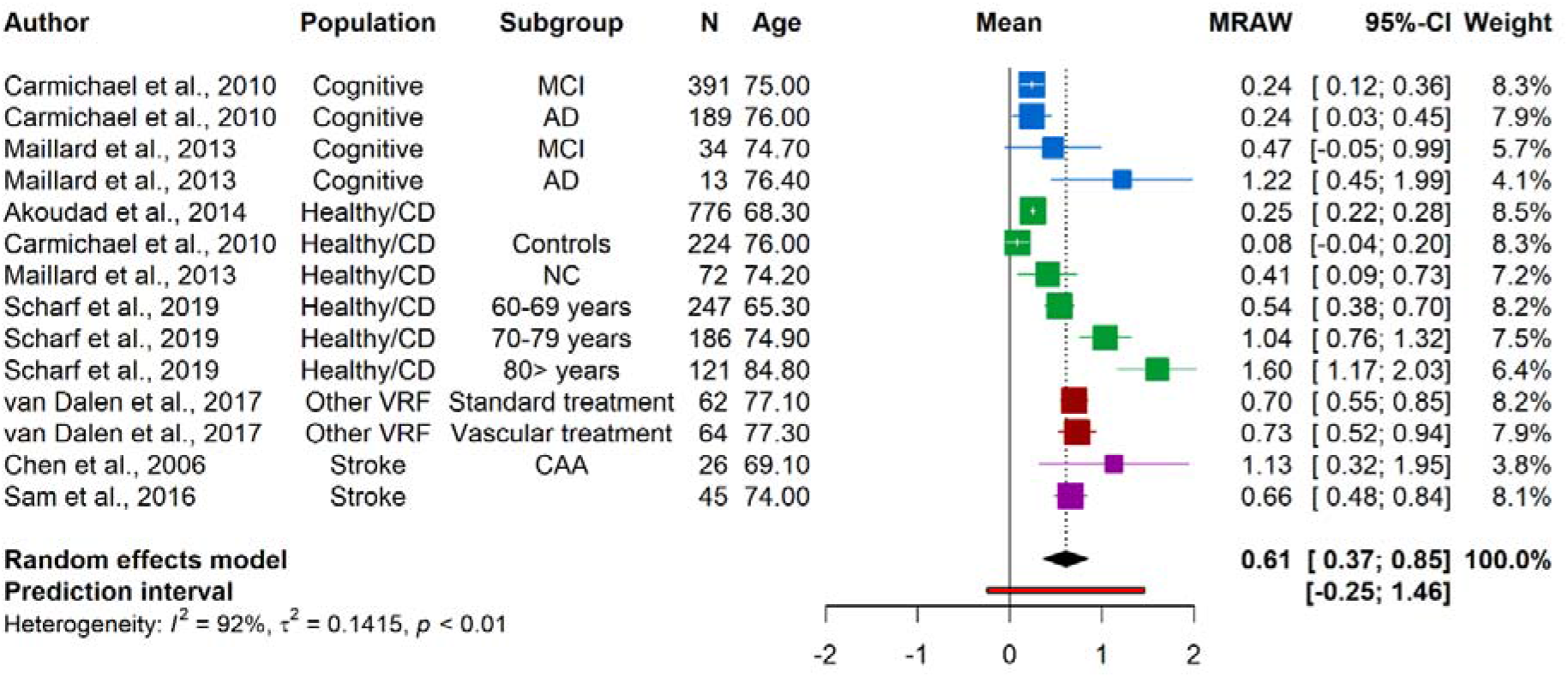
Random-effects meta-analysis of mean WMH volume in mL/year. Squares represent means and bars the 95% CI. Note: AD: Alzheimer’s disease; CAA: Cerebral amyloid angiopathy; Healthy/CD: healthy/community-dwelling; MCI: Mild cognitive impairment; MRAW: raw means; NC: normal controls; VRF: Vascular risk factors

### Subgroup analyses

For explorative purposes, we examined WMH change per population type and, when possible, by age and time lapse between scans.

#### Population

We performed meta-analyses per population group for all three ways of reporting WMH change (mL, %ICV, mL/year). Where studies reported treatment groups, control or case groups, these subgroups are reported. Not all population groups were available for %ICV and mL/year.

WMH raw volume increased by 1.78 mL (95% CI 0.83, 2.73; PI -2.18, 5.75; eFigure 1) in healthy and community-dwelling population over 2.9±1.3 years (range 1 – 5.3).^10, 11, 13-16, 19-21, 23-26, 28, 29, 31, 33^ WMH %ICV increased by 0.26 %ICV (95% CI 0.09, 0.43; PI -0.20, 0.71; eFigure 2), over 1.9 – 6.7 years^35-37, 40^ and the annual rate by 0.62 mL/year (95% CI 0.02, 1.22; PI -1.03, 2.27; eFigure 3).^41, 42, 44, 46^

In people with memory complaints^22^ and dementia^10, 24^ WMH increased by mean 1.17 mL (95% CI 0.40, 1.94; PI -0.87, 3.20; eFigure 4). Among mild cognitive impairment and Alzheimer’s dementia groups, WMH increased by 0.27mL/year (95% CI 0.02, 0.51; PI -0.07,0.60; eFigure 5).^42, 44^

In people with depression, WMH volume increased by 1.19 mL (95% CI -1.81, 4.18; PI -6.90, 9.27; eFigure 6).^18, 27^ Within the depression populations, a group with incident dementia at follow up^27^ presented the largest mean WMH volume increase with 4.52mL (95% CI 2.25, 6.79), while the raw mean volume changes for the other depression groups range from 0.08-1.10 mL.

Among populations recruited because of vascular risk factors (VRF), i.e., participants with diabetes,^12, 35^ SVD,^4, 17^ multiple risk factors,^32, 47^ hypertension,^13, 38^ and vascular disease or high risk of vascular disease,^30^ WMH volume also increased on average, including increases of 2.02mL (95% CI 0.95, 3.09; PI -1.19, 5.23; eFigure 7),^4, 12, 13, 17, 30, 32^ 0.30%ICV (95% CI -0.14, 0.74; PI -2.22, 2.81; eFigure 8),^35, 38^, and 0.71 mL/year (95% CI 0.53, 0.89; PI not calculable; eFigure 9).^47^

Data on stroke patients was available for WMH volume in mL, increase of 2.46 mL (95% CI - 0.21, 5.12; PI -4.50, 9.41; eFigure 10),^5, 28, 34^ and mL per year, an increase of 0.72 mL/year(95% CI -1.32, 2.76; PI not calculable; eFigure 11).^43, 45^

#### Time between scans

Most studies had a follow up time between scans of around two years. The 27 papers reporting raw volume in mL had a follow-up time of 2.7±1.65 years (median=2, range 0.25 – 8.7). Longer follow up times between scans appeared associated with larger WMH increase in mL (Figure 5). In a similar bubble plot of mean WMH change as %ICV (3.5 ± 1.54 years, median=3; range 1.9 – 6.7), there is no clear relation between longer time between scans and larger WMH change (Figure 6) but there were far fewer studies.

**Figure 5.**
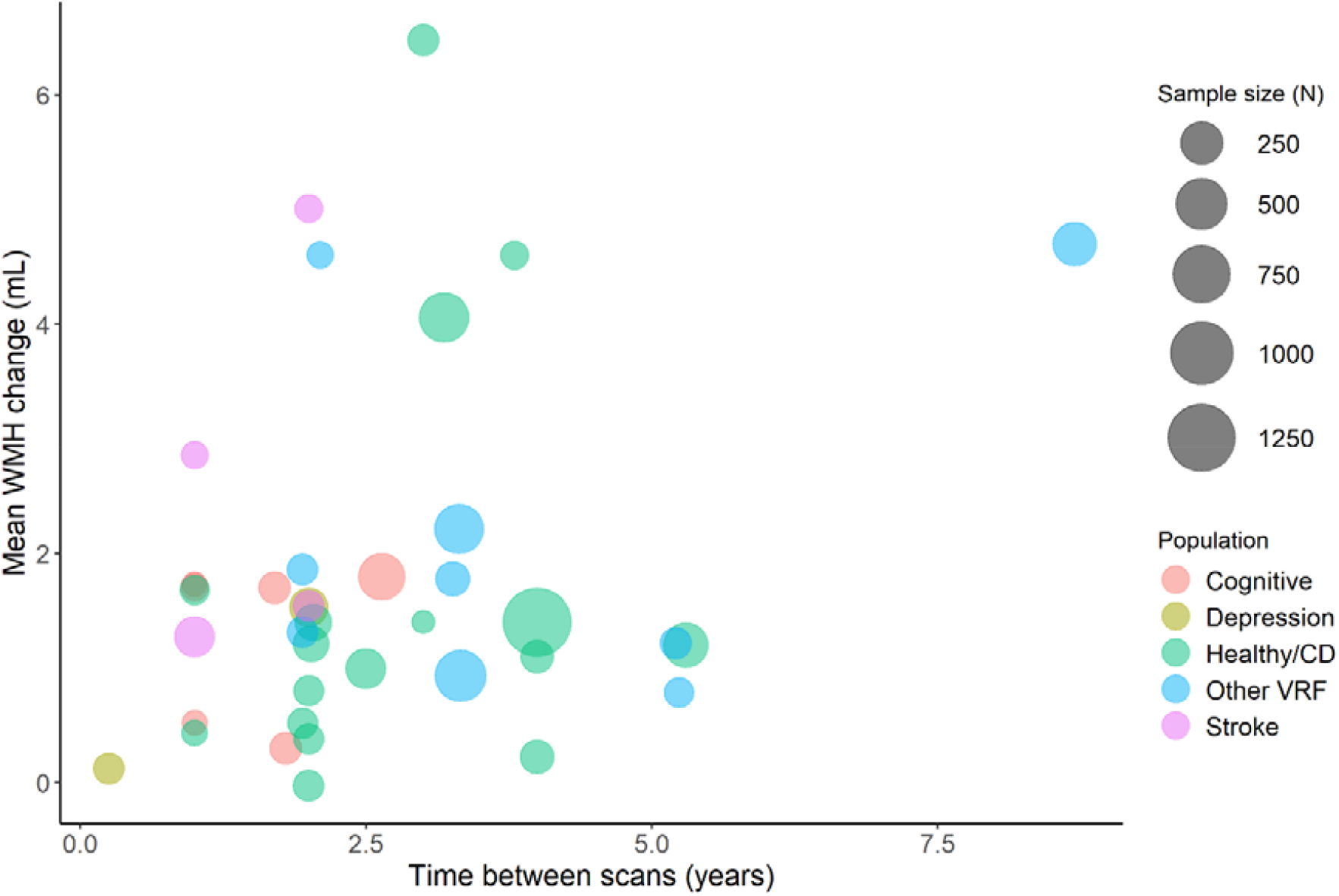
Bubble plot of mean WMH change (mL), in individual studies, related to time between scans (years). Points in the plot are scaled by sample size, and the colour of points refer to population group.

**Figure 6.**
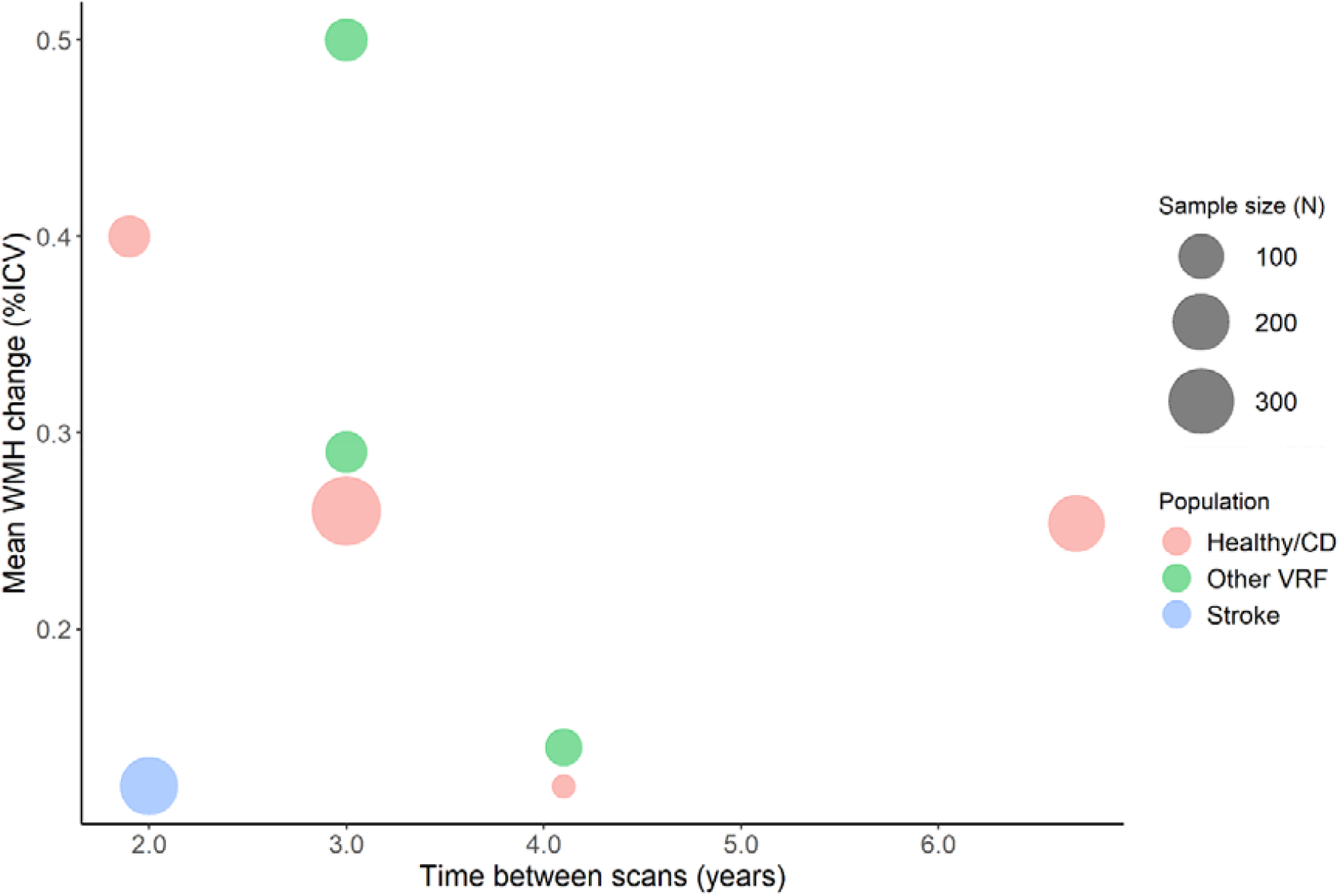
Bubble plot of mean WMH change (%ICV), in individual studies, related to time between scans (years). Points in plot are scaled by sample size and colour of points refer to population group.

#### Age

Patterns in the bubble plots of WMH change versus mean age at baseline suggest that WMH volume change increases at older ages across WMH in mL (Figure 7), %ICV (eFigure 12) and mL/year (eFigure 13), with a younger mean age at baseline generally corresponding to smaller WMH change over time.

**Figure 7.**
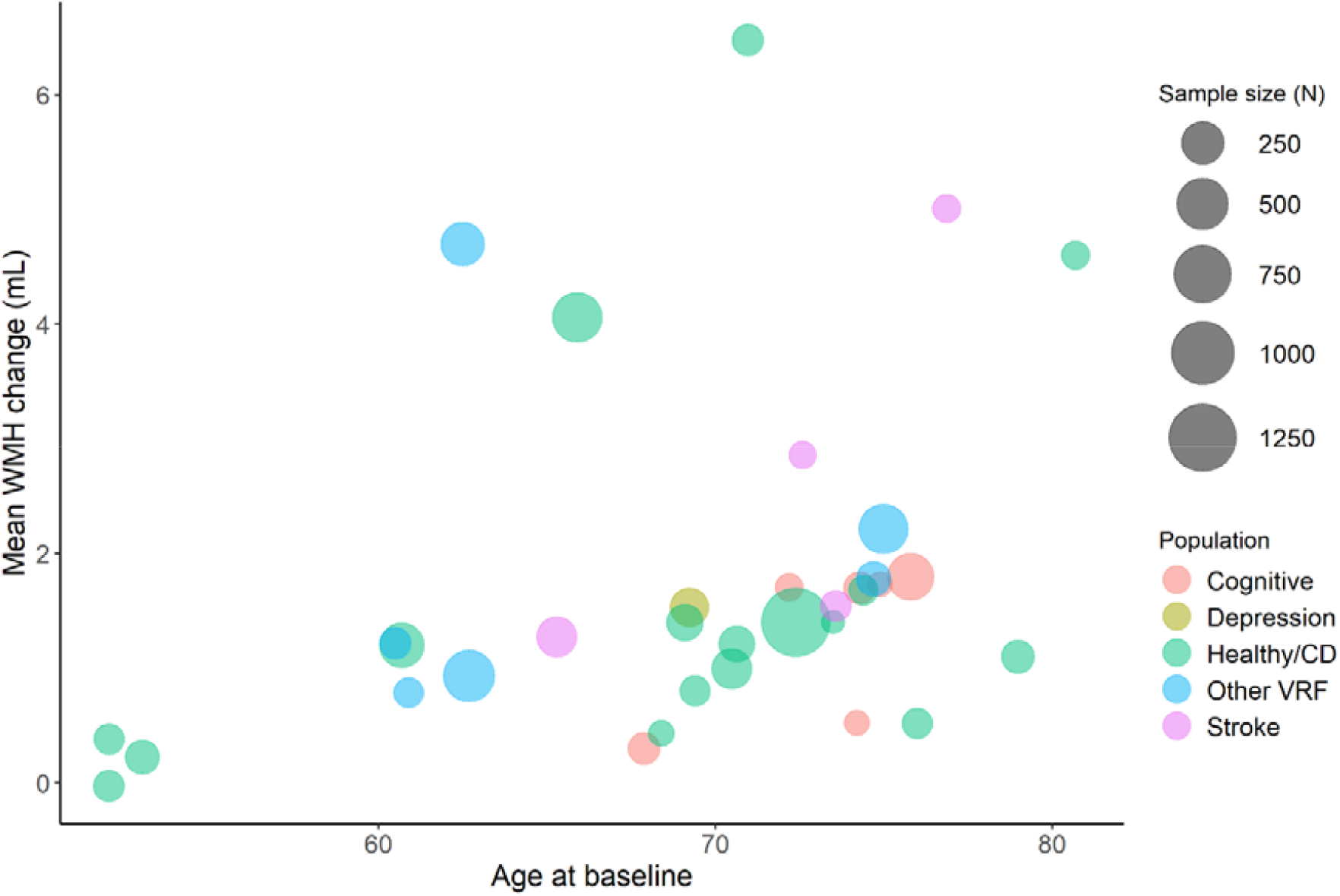
Bubble plot of mean WMH change (mL) related to age at baseline (years). Points in plot are scaled by sample size and colour of points refer to population group.

### WMH regression

Thirteen of the 39 papers (eTables 3-5) included in the meta-analysis mentioned WMH volume regression, of which WMH volume regression was discussed by 8/13 papers,^4, 5, 11, 16, 24, 26, 45, 46^ but only 7/8 papers^5, 10, 16, 22, 23, 25, 28^ provided data. One paper^46^ did not mention regression but showed WMH regression in a figure with individual trajectories.

WMH regression was found in healthy/community-dwelling participants (in ∼34%^11^ and 17%^26^) and in stroke participants (4%^45^ and 37%^5^). Areas of WMH shrinkage were found in AD participants with high SVD load, with less WMH regression in cognitively normal controls who had less WMH at baseline than the patients with AD and high SVD load.^24^ Over 9-years follow-up^4^ (n=276), one participant (0.4%) showed net WMH regression, while 9% showed regression in the first follow-up period (2006-2011) and 2% in the second period (2011-2015). Factors associated with WMH regression were not found.^48^ Observations of WMH regression from papers not included in meta-analysis (eTable 6) can be found in eResult1, and associations with WMH change in eResult2.

## Discussion

Our meta-analyses suggest that although WMH volumes increase on average, WMH volumes also regress, with WMH volume regression occurring explicitly in up to 1/3 of participants. The prediction intervals of the main analyses of WMH change capture the extent of WMH volume regression (−1.24 mL; -0.06 %ICV; -0.25 mL/year) and increase (4.73 mL; 0.56 %ICV; 1.46 mL/year). We show that WMH regression can occur in all typical populations affected by SVD, greater WMH volume change might occur at older age, and WMH regression might occur over a wide range of follow-up times.

As WMH progression is the main focus of most papers and WMH regression is regarded as an accidental finding or even an ‘error’, the underlying mechanisms of WMH regression are unknown, or whether regression represents improvement in tissue health and translates to a positive effect on clinical outcomes.^48^ However, some evidence suggests that regression is linked to less cognitive decline, recurrent stroke or dependency.^49^ Furthermore, if patients with less WMH progression have less cognitive impairment than those with more WMH progression, then there is reason to think that WMH regression might translate to even better outcomes.

The papers included had several limitations. First, methodologies to measure and report WMH volume change, i.e. mL, %ICV, or mL/year, varied and complicated the comparison of volume change, and means that the findings should be interpreted with caution. Although there are many methods to assess WMH volume, including artificial intelligence approaches, there is little cross-validation or standardisation. Furthermore, methods specifically addressing WMH volume change that account for registration steps are only beginning to emerge.^50^ Second, papers that report WMH volume change as an annualised rate might represent a bias by assuming that change is linear. We did not find a clear relation between longer follow-up times and larger WMH volume change but this may reflect the heterogeneity of studies, populations and follow-up times. A study with a 9-year follow-up period^4^ shows that WMH volumes can both increase and decrease within the 9 years and that more people had WMH regression in the first five years than in the second part. Thus, studies reporting annualized rates should also report the absolute change over time at the final time point. Our analyses were limited by not being able to examine factors related to regression since too few papers examined these. For the same reason, we were unable to assess the effect of interventions on WMH volume regression.

The strengths of our review include a comprehensive literature search of WMH volume change using different measurements of WMH volume; subgroup exploratory analyses into differences between populations, age and follow-up times; and a good geographical coverage of included studies. The review uses prediction intervals to show least and most change, rather than only confidence intervals which focus on the mean and thus obscure the true limits of change. Hence the review demonstrates the range of inter-individual differences in WMH progression volume that may have been overlooked through the tendency in previous studies to focus solely on WMH progression.

Future studies should examine WMH change including the possibility of WMH regression and investigate WMH change over a long period of time. The median follow-up times in our main analyses were 2 years (mL) and 3 years (%ICV), providing little evidence over longer follow-times^2^. Longer follow-up times and scans at multiple time-points would provide more information on trajectories of volume change and dynamics of WMH. Additionally, examination of possible pathological, imaging and clinical factors related to WMH regression is vital as the exact underlying mechanisms and clinical consequences are unknown. In conclusion, our results indicate that WMH volumes can regress over time in diverse populations, while net WMH volume might progress. However, little is known about underlying mechanisms of WMH volume regression while WMH changes might represent an opportunity to develop new interventions and delay the progression of WMH and its devastating clinical consequences.

## Supporting information

PRISMA checklists

## Data Availability

Data produced in the present study are available upon reasonable request

## Disclosure

None

## Funding

ACCJ was funded by the Alzheimer’s Society (ref 486 (AS-CP-18b-001), University of Edinburgh College of Medicine and Veterinary Medicine and the UK Dementia Research Institute which receives funding from UK DRI Ltd funded by the UK Medical Research Council, Alzheimer’s Society and Alzheimer’s Research UK. CA is funded by UK Dementia Research Institute as above, the Mexican National Council of Science and Technology (CONACYT) and the Anne Rowling Regenerative Neurology Clinic. TR and MH were funded by the Wellcome Trust through the Translational Neurosciences PhD program. SMM was funded by Age UK. FND acknowledges support from The Stroke Association, Garfield Weston Foundation, NHS Research Scotland and the Agnes Parry Endowment at the University of Edinburgh. JMW acknowledges support from the Fondation Leducq Transatlantic Network of Excellence for the Study of Perivascular Spaces in Small Vessel Disease (16 CVD 05) and the UK Dementia Research Institute as above.

## Notes

### Competing Interest Statement

The authors have declared no competing interest.

