## Supplemental material for "Longitudinal changes of white matter hyperintensities in sporadic small vessel disease: a systematic review and meta-analysis"

**Contents Supplemental Material**

eAppendix 1: Search strategy

eTable 1: Data extraction form

eTable 2: Risk of bias assessment

eTable 3: Summary of included papers – WMH volume in mL

eTable 4: Summary of included papers – WMH volume as %ICV

eTable 5: Summary of included papers – WMH volume as mL per year

eTable 6: Summary of relevant papers without useable WMH change data

eResult 1: WMH regression

eResult 2: Summary of WMH changes

eFigures 1 – 13

eReferences

**eAppendix 1: Search Strategy**

**Database: Embase 1974 to 28 January 2022**

Embase Search strategy 2 WMH –

1. ((WM OR WMH OR white matter OR white matter hyperintensit*) adj3 (shr?nk* OR reduc* OR recover* OR regress* OR diminish* OR decrease* OR lower)).tw.

2. ((WM OR WMH OR white matter OR white matter hyperintensit*) adj3 (gr?w OR enlarge* OR worse* OR progress* OR increas* OR change* OR high*)).tw.

3. 1 OR 2

4. Leukoaraiosis/

5. (((WM OR white matter) adj3 (hyperintens* OR lesion* OR abnormalit* OR disease OR damage OR abnormal* OR disease*)) OR leukoarasios* OR WMH).tw.

6. 4 OR 5

7. Cerebral small vessel diseases/

8. (Cerebral small vessel disease OR SVD OR CSVD OR ((disease adj3 (small adj2 (vessel* OR vasculature OR arter* OR capillar*))) OR microvasculature)).tw.

9. 7 OR 8

10. exp Cohort studies/ OR disease progression/ OR Interrupted Time Series Analysis/ OR Controlled Before-After Studies/ OR exp Clinical Study/

11. (follow-up OR longitudinal* OR over time OR follow* up OR retrospective* OR prospective* OR disease progress* OR cohort study OR outcome OR evolution OR at baseline).tw.

12. 10 OR 11

13. exp Magnetic resonance imaging/

14. (MRI OR magnetic resonance OR MR OR image ajd1 analys* OR T2 OR FLAIR OR fluid attenuation inversion recovery OR DWI OR diffusion weighted).tw.

15. 13 OR 14

16. 3 and 6 and 9 and 12 and 15

**Database: Ovid MEDLINE (R) 1946 to 28 January 2022**

Search Strategy:

1. ((WM OR WMH OR white matter OR white matter hyperintensit*) adj3 (shr?nk* OR reduc* OR recover* OR regress* OR diminish* OR decrease* OR lower)).tw.

2. ((WM OR WMH OR white matter OR white matter hyperintensit*) adj3 (gr?w OR enlarge* OR worse* OR progress* OR increas* OR change* OR high*)).tw.

3. 1 OR 2

4. Leukoaraiosis/

5. (((WM OR white matter) adj3 (hyperintens* OR lesion* OR abnormalit* OR disease OR damage OR abnormal* OR disease*)) OR leukoarasios* OR WMH).tw.

6. 4 OR 5

7. Cerebral small vessel diseases/

8. (Cerebral small vessel disease OR SVD OR CSVD OR ((disease adj3 (small adj2 (vessel* OR vasculature OR arter* OR capillar*))) OR microvasculature)).tw.

9. 7 OR 8

10. exp Cohort studies/ OR disease progression/ OR Interrupted Time Series Analysis/ OR Controlled Before-After Studies/ OR exp Clinical Study/

11. (follow-up OR longitudinal* OR over time OR follow* up OR retrospective* OR prospective* OR disease progress* OR cohort study OR outcome OR evolution OR at baseline).tw.

12. 10 OR 11

13. exp Magnetic resonance imaging/

14. (MRI OR magnetic resonance OR MR OR image ajd1 analys* OR T2 OR FLAIR OR fluid attenuation inversion recovery OR DWI OR diffusion weighted).tw.

15. 13 OR 14

16. 3 and 6 and 9 and 12 and 15

**eTable 1: Data extraction form**

| Article name: | |
| --- | --- |
| First Author |  |
| Year |  |
| Study name |  |
| Study registration |  |
| Country |  |
| Population |  |
| Study Design | Cohort □ Case-control □ Randomized trial □  Non-randomized trial □ |
| Control group(s) (if any) |  |
| Sample size baseline |  |
| Sample size follow-up |  |
| Duration of follow-up |  |
| **Demographic variables** | |
| Mean age ± SD (or median/IQR/range if no mean) at baseline (and follow-up if available) |  |
| Hypertension, N (%) |  |
| Diabetes, N (%) |  |
| Hypercholesterolemia, N (%) |  |
| History of smoking, N (%) |  |
| Body mass index, mean (SD) |  |
| Education, mean (SD) |  |
| MMSE/MoCA, mean (SD) |  |
| Comments: interventions if trial |  |
| **Imaging variables** | |
| Scanner | 1.5T □ 3T □ 7T □ |
| MRI sequence used for WMH volume |  |
| WMH unit of volume, e.g. mL, cc, cm3, %ICV |  |
| WMH volume at baseline  (mean, SD or median, range, IQR, 95% CI if no mean and SD) |  |
| WMH volume at follow up  (mean, SD or median, range, IQR, 95% CI if no mean and SD) |  |
| WMH volume change  (mean, SD or median, range, IQR, 95% CI if no mean and SD) |  |
| Definition of change |  |
| Volume corrected for ICV or TBV | No □ ICV □ TBV □ |
| Methods of quantifying WMH volumes |  |
| Slice thickness |  |
| Predictors of WMH change (if assessed) |  |
| Comments |  |
| **Risk of Bias Assessment for Non-randomized Studies (RoBANS)** | |
| 1. The selection of participants | Low risk □ High risk □ Unclear risk □ |
| 2. Confounding variables | Low risk □ High risk □ Unclear risk □ |
| 3. Measurement of exposure | Low risk □ High risk □ Unclear risk □ |
| 4. Blinding of outcome assessments | Low risk □ High risk □ Unclear risk □ |
| 5. Incomplete outcome data | Low risk □ High risk □ Unclear risk □ |
| 6. Selective outcome reporting | Low risk □ High risk □ Unclear risk □ |

**eTable 2: Risk of bias assessment**

|  | **Selection of participants** | **Confounding variables** | **Measurement of exposure** | **Blinding of outcome assessments** | **Incomplete outcome data** | **Selective outcome reporting** |
| --- | --- | --- | --- | --- | --- | --- |
| Akoudad et al., 2014^1^ | LOW | LOW | LOW | LOW | LOW | LOW |
| Burton, 2006^2^ | LOW | LOW | LOW | UNCLEAR | UNCLEAR | LOW |
| Callisaya, 2015^3^ | LOW | LOW | UNCLEAR | LOW | LOW | LOW |
| Carmichael, 2010^4^ | LOW | LOW | LOW | UNCLEAR | HIGH | LOW |
| Chen, 2006^5^ | LOW | LOW | LOW | LOW | LOW | LOW |
| De Bresser, 2010^6^ | LOW | LOW | LOW | UNCLEAR | LOW | LOW |
| De Havenon, 2019^7^ | LOW | LOW | LOW | UNCLEAR | LOW | LOW |
| Dickie, 2016 ^8^ | LOW | LOW | LOW | UNCLEAR | LOW | LOW |
| Firbank, 2007 ^9^ | LOW | LOW | LOW | LOW | LOW | LOW |
| Garde, 2005^10^ | LOW | LOW | UNCLEAR | LOW | LOW | LOW |
| Godin, 2009^11^ | LOW | LOW | LOW | LOW | LOW | LOW |
| Gopalan, 2014^12^ | LOW | LOW | LOW | LOW | LOW | LOW |
| Gouw, 2008^13^ | LOW | LOW | LOW | LOW | UNCLEAR | LOW |
| Khalaf, 2015^14^ | LOW | LOW | LOW | UNCLEAR | UNCLEAR | LOW |
| Kneihsl, 2020^15^ | LOW | LOW | LOW | LOW | UNCLEAR | LOW |
| Lee, 2017^16^ | LOW | LOW | LOW | LOW | LOW | LOW |
| Maillard, 2013^17^ | LOW | LOW | LOW | UNCLEAR | LOW | LOW |
| Marquine, 2010^18^ | LOW | LOW | LOW | UNCLEAR | UNCLEAR | LOW |
| Moon, 2018^19^ | LOW | LOW | LOW | LOW | UNCLEAR | LOW |
| Moscufo, 2012^20^ | UNCLEAR | LOW | LOW | UNCLEAR | LOW | LOW |
| Nebes, 2002^21^ | UNCLEAR | LOW | LOW | LOW | LOW | LOW |
| Ramirez, 2016^22^ | LOW | LOW | LOW | UNCLEAR | UNCLEAR | LOW |
| Raz, 2013^23^ | LOW | LOW | LOW | LOW | UNCLEAR | LOW |
| Sachdev, 2007^24^ | LOW | HIGH | LOW | UNCLEAR | LOW | LOW |
| Sam, 2016^25^ | LOW | LOW | LOW | UNCLEAR | UNCLEAR | LOW |
| Scharf, 2019^26^ | LOW | LOW | LOW | UNCLEAR | LOW | LOW |
| Steffens, 2007^27^ | LOW | LOW | LOW | UNCLEAR | LOW | LOW |
| Switzer, 2016^28^ | UNCLEAR | HIGH | LOW | LOW | LOW | LOW |
| Taylor, 2003^29^ | LOW | LOW | LOW | UNCLEAR | LOW | LOW |
| Van Dalen, 2017^30^ | UNCLEAR | LOW | LOW | LOW | LOW | LOW |
| Van Elderen, 2010^31^ | LOW | LOW | LOW | UNCLEAR | LOW | LOW |
| Van Leijsen, 2017^32^ | LOW | LOW | LOW | LOW | LOW | LOW |
| Van Sloten, 2015^33^ | LOW | LOW | LOW | LOW | LOW | LOW |
| Wardlaw, 2017^34^ | LOW | LOW | LOW | LOW | LOW | LOW |
| White, 2019^35^ | LOW | LOW | LOW | LOW | LOW | LOW |
| Whitman, 2001^36^ | UNCLEAR | LOW | LOW | LOW | LOW | LOW |
| Wyss, 2019^37^ | LOW | LOW | LOW | LOW | LOW | LOW |
| Xia, 2020^38^ | LOW | LOW | LOW | LOW | LOW | LOW |
| Xiong, 2014^39^ | LOW | LOW | LOW | LOW | LOW | LOW |
| Yoon, 2017^40^ | LOW | LOW | UNCLEAR | UNCLEAR | LOW | LOW |

Red = high risk of bias, Green = low risk of bias, Yellow = unclear according to the review authors

**eTable 3: Summary of included papers – WMH volume in mL**

| **Author, year** | **Study design** | **Population** | **Sub-group** | **N (^a^)** | **Mean age (SD)^a^** | **Sex, male (%)^a^** | **Mean time between scans, years** | **WMH measure, sequence, scanner** | **Predictors**  **of WMH change** | **Baseline WMH volume, mean** **± SD** | **WMH change,**  **mean ± SD** | **WMH regression mentioned by authors** |
| --- | --- | --- | --- | --- | --- | --- | --- | --- | --- | --- | --- | --- |
| Burton et al., 2006^2^ | Case-control | Healthy/CD | Controls | 33 (33) | 74.4 (6.3) | 19 (57.6) | 1 | mL, FLAIR, 1.5T | Progression: Baseline WMH | 5, 2-13 mL  (geometric mean, 1 SD range) | 1.68 ± 4.02 mL  Median, range^c^:  0.24, -1.7 - 15 mL | no |
| Burton et al., 2006^2^ | Case-control | Cognitive | AD | 23 (23) | 78.2 (4.6) | 13 (56.5) | 1 | mL, FLAIR, 1.5T | Progression: Baseline WMH | 14, 5-41  (geometric mean, 1 SD range) | 1.70 ± 3.55 mL  Median, range^c^:  0.77, -2.7 - 11 mL | no |
| Burton et al., 2006^2^ | Case-control | Cognitive | DLB | 14 (14) | 74.2 (8.1) | 8 (57.1) | 1 | mL, FLAIR, 1.5T | Progression: Baseline WMH | 5, 2-9  (geometric mean, 1 SD range) | 0.52 ± 1.14 mL  Median, range^c^:  0.29, -1.0 – 2.9 mL | no |
| Burton et al., 2006^2^ | Case-control | Cognitive | PDD | 13 (13) | 74.9 (3.5) | 8 (61.5) | 1 | mL, FLAIR, 1.5T | Progression: Baseline WMH | 6, 2-13  (geometric mean, 1 SD range) | 1.73 ± 2.48 mL  Median, range^c^:  0.78, -0.8 – 7.5 mL | no |
| Callisaya et al., 2015^3^ | Cohort | Healthy/CD | - | 187 (353) | 70.4 (6.5) | 97 (27) | 2.5 | mL, FLAIR, T1, T2. 1.5T | - | 6.18 ± 6.60 mL | 0.99 ± 2.60 mL | yes; dismissed as measurement error |
| de Havenon et al., 2019^7^ | Randomized trial | Other VRF | Cohort 1. DM II, Intensive and standard glycaemic arms | 502 (632) | 62.7 (5.7) | 270 (42.7) | 3.33 | cm^3^, FLAIR, - | Progression: Higher systolic blood pressure.  Reduced WMH progression: Intensive blood pressure control treatment | 1.76 ± 2.50 mL | 0.93 ± 1.2 mL | no |
| Firbank et al., 2007^9^ | Randomized trial | Healthy/CD | Normotensive | 41 (41) | 76 (4) | 28 (68.3) | 1.95^b^ | mL, FLAIR, 1.5T | Progression: Deep WMH at baseline, diastolic BP between scans, baseline systolic BP | 7.70 ± 3.89 mL^f^ | 0.52 ± 1.98 mL^f^ | no |
| Firbank et al., 2007^9^ | Randomized trial | Other VRF | Hypertensive – placebo | 47 (47) | - | - | 1.95^b^ | mL, FLAIR, 1.5T | Progression: deep WMH at baseline, diastolic BP between scans, baseline systolic BP | 12.40 ± 7.18 mL^f^ | 1.86 ± 3.25 mL^f^ | no |
| Firbank et al., 2007^9^ | Randomized trial | Other VRF | Hypertensive – drug (candesartan) | 45 (45) | - | - | 1.95^b^ | mL, FLAIR, 1.5T | Progression: deep WMH at baseline, diastolic BP between scans, baseline systolic BP | 11.3 ± 6.73 mL^f^ | 1.31 ± 3.24 mL^f^ | no |
| Garde et al., 2005^10^ | Cohort | Healthy/CD | - | 26 | 80.7 (0.4) | 16 (62) | 3.8 | cm^3,^ T2, proton density,  1.5T | - | 4.7, 0 – 44.2 mL  Median, range | 6.5 ± 5.2 mL  Median, range^c^:  2.6, 0.1 - 20.7 mL | no |
| Godin et al., 2009^11^ | Cohort | Healthy/CD | - | 1319 (1779) | 79.92 (5.68) | 703 (39.5) | 4 | cm^3^, T1, T2. 1.5T | Progression: Apolipoprotein E genotype (ε4ε4) | NA | 1.4 ± 2.8 mL | no |
| Gopalan et al., 2014^12^ | Randomized trial | Healthy/CD | Placebo | 42 (59) | 52 (8.2) | 21 (35.6) | 2 | mm^3^, T1, T2, FLAIR, 1.5T | Reduced progression: Mixed tocotrienols | 1633 ± 232 mm^3^  Mean ± SEM | 0.38 ± 0.94 mL  Mean, SEM: 380, 123 mm^3^ | no; dismissed as negligible change |
| Gopalan et al., 2014^12^ | Randomized trial | Healthy/CD | T3 | 46 (62) | 52 (8.8) | 27 (43.5) | 2 | mm^3^, T1, T2, FLAIR, 1.5T | Reduced progression: Mixed tocotrienols | 1523 ± 182 mm^3^  Mean ± SEM | -0.03 ± 0.94 mL  Mean, SEM^c^: -29, 119 mm^3^ | no; dismissed as negligible change |
| Gouw et al., 2008^13^ | Cohort | Other VRF | - | 20 (20) | - | - | 2.1 | mL, FLAIR, - | - | 24.6 ± 27.9 mL | 4.6 ± 5.1 mL | no |
| Khalaf et al., 2015^14^ | Case-control (using participants from trials) | Depression | Total | 47 (47) | - | 15 (31.9) | 0.25^d^ | cm^3^, FLAIR, T2, 3T | Progression: Mainly driven by non-remitters | 0.56 ± 0.77 mL | 0.12 ± 0.35 mL | no |
| Khalaf et al., 2015^14^ | Case-control (using participants from trials) | Depression | Remitters | 23 (23) | 68.0 (5.7) | 9 (39.1) | 0.25^d^ | cm^3^, FLAIR, T2, 3T | Progression: Mainly driven by non-remitters | 0.41 ± 0.30 mL | 0.08 ± 0.34 mL | no |
| Khalaf et al., 2015^14^ | Case-control (using participants from trials) | Depression | Non-remitters | 24 (24) | 66.8 (5.7) | 6 (25) | 0.25^d^ | cm^3^, FLAIR, T2, 3T | Progression: Mainly driven by non-remitters | 0.71 ± 1.03 mL | 0.16 ± 0.37 mL | no |
| Kneihsl et al., 2020^15^ | Cohort | Healthy/CD | - | 293 (491) | 60.7 (6.9) | 253 (51.5) | 5.3 | cm^3^, T2 and FLAIR. 1.5T | Progression: Arterial hypertension | NA | 1.2 ± 3.3 mL | no |
| Lee et al., 2017^16^ | Cohort | Healthy/CD | - | 450 (450) | 65.9 (8.2) | 226 (50.2) | 3.2^d^ | mL, FLAIR, 1.5T or 3T | Progression: Resistance index ratio along the middle cerebral artery | 9.66 ± 14.54 mL  3.72,  1.25 – 13.87  Median, IQR | 4.06 ± 7.35 mL  1.17,  0.09 – 4.93  Median, IQR | no |
| Marquine et al., 2010^18^ | Cohort | Healthy/CD | - | 110 (110) | 70.7 (5.6) | 30 (27) | 2^b^ | mL, dual-echo fast spin-echo. 1.5T | - | 4.92 ± 7.58 mL | 1.21 ± 2.30 mL | no |
| Moon et al., 2018^19^ | Randomized trial | Cognitive | Total | 358 (458) | 75.5 (3.9) | 130 (36.3) | 2.6^b^ | cm3, FLAIR, - | - | 9.8,  6.6 – 16.3 cm^3^  Median, IQR | 1.77 ± 3.20 mL  Median, IQR^c^:  1.2, -0.1 – 4.2 | no |
| Nebes et al., 2002^21^ | Case-control | Healthy/CD | Control | 12 (12) | 73.5 (4.4) | - | 3 | mm3, FLAIR, 1.5T | - | 5315 ± 9863 mm^3^ | 1.40 ± 2.61 mL  Median, 95% CI^c^: 1404,  -75 – 2883 | no |
| Ramirez et al., 2016^22^ | Case-control | Healthy/CD | Normal controls | 44 (44) | 69.4 (7.0) | 19 (43.2) | 2 | cc, T1, PD/T2 weighted axial dual echo spin echo, 1.5T | - | 4.4 ± 5.6 mL | 0.8 ± 2 mL | yes |
| Ramirez et al., 2016^22^ | Case-control | Cognitive impairment | AD patients with low SVD burden | 56 (56) | 67.9 (8.0) | 28 (50) | 1.8 | cc, T1, PD/T2 weighted axial dual echo spin echo, 1.5T | - | 1.3 ± 0.9 mL | 0.3 ± 1.1 mL | yes |
| Ramirez et al., 2016^22^ | Case-control | Cognitive impairment | AD patients with high SVD burden | 57 (57) | 74.3 (8.3) | 22 (38.6) | 1.7 | cc, T1, PD/T2 weighted axial dual echo spin echo, 1.5T | - | 14.0 ± 13.7 mL | 1.7 ± 5.1 mL | yes |
| Raz et al., 2013^23^ | Randomized trial | Healthy/CD | - | 79 (95) | 53 (51-54 IQR) | 0 (0) | 4^d^ | mm3, FLAIR, MPRAGE, 1.5T | Progression: fibrinogen receptor complex on platelets, platelet-derived microvesicles, and thrombogenic microvesicles | 1909,  15.7 – 2655 mm^3^  Median, IQR | 0.22 ± 0.53 mL  Median, IQR^c^:  155.4, -92.13 – 599.4 mm^3^ | no |
| Sachdev et al.,2007 ^24^ | Cohort | Healthy/CD | - | 51 (80; BL data reported for 51) | 71.0 (5.9) | 26 (51) | 3 | Cc, FLAIR, T1, 1.5T | Progression: WMH at baseline | 16.38 ± 13.79 mL | 6.48 ± 7.64 mL | yes |
| Steffens et al., 2007^27^ | Cohort | Depression | Total | 161 (161) | 69.2 (7.1) | 61 (38) | 2 | mL, dual-echo fast spin-echo. 1.5T | - | 6.54 ± 10.78 mL | 1.53 ± 3.53 mL | no |
| Steffens et al., 2007^27^ | Cohort | Depression | Incident Dementia | 20 (20) | 76.1 (1.2) | 3 (15) | 2 | mL, dual-echo fast spin-echo. 1.5T | - | 11.82 ± 15.80 mL | 4.52 ± 5.19 mL | no |
| Steffens et al., 2007^27^ | Cohort | Depression | No incident Dementia | 141 (141) | 68.3 (0.6) | 58 (41) | 2 | mL, dual-echo fast spin-echo. 1.5T | - | 5.80 ± 9.72 mL | 1.1 ± 3.02 mL | no |
| Switzer et al., 2016^28^ | Case-control | Healthy/CD | Controls | 16 (16) | 68.4 (5.9) | 7 (43.8) | 1 | mL, FLAIR, 3T | - | 2.34,  2.71 – 5.27 mL  Median, IQR | 0.42 ± 1.01 mL  Median, IQR^c^: 0.33, -0.15 – 1.09 mL | no |
| Switzer et al., 2016^28^ | Case-control | Stroke | CAA | 22 (22) | 72.6 (6.9) | 14 (63.6) | 1 | mL, FLAIR, 3T | - | 24.8,  20.77 – 44.01 mL  Median, IQR | 2.75 ± 6.22 mL    Median, IQR^c^: 1.36,  -0.48 – 7.37 mL | no |
| Taylor et al., 2003^29^ | Cohort | Healthy/CD | - | 117 (117) | 69.1 (6.20) | 26 (21.9) | 2.0^b^ | cc, dual-echo fast spin-echo. 1.5T | Progression: Age and self-reported diabetes.  Less WMH change: Symptoms of cardiac disease | 4.91 ± 7.01 mL  Range: 0.41- 49.98 | 1.4 ± 3.04 mL | no |
| van Elderen et al., 2010^31^ | Case-control (substudy randomized trial) | Other VRF | DM | 89 (89) | 74.7 (3.1) | 53 (60) | 3.26^e^ | cc, T2, FLAIR, 1.5T | - | 4.19 ± 0.82 mL | 1.78 ± 0.29 mL | no |
| van Elderen et al., 2010^31^ | Case-control (substudy randomized trial) | Other VRF | Controls | 438 (438) | 75.0 (3.2) | 244 (56) | 3.32^e^ | cc, T2, FLAIR, 1.5T | - | 5.34 ± 0.48 mL | 2.21 ± 0.17 mL | no |
| van Leijsen et al., 2017^32^ | Cohort | Other VRF | Overall change (2006-2015) | 276 (276) | 62.5 (7.7) | 163 (59.1) | 8.7 | mL, FLAIR, 1.5T | Progression: Age | 5.8 ± 9.5 mL  Median, IQR 2.3, 0.8–6.1 | 4.7 ± 6.6 mL  Median, IQR 2.1, 0.95–5.5 | yes |
| Van Sloten et al., 2015^33^ | Cohort | Healthy/CD | - | 1949 (1949) | 74.6 (4.6) | 846 (43.4) | 5.2 | mL, proton desnity/T2 weighted fast spin-echo, FLAIR, 1.5T | - | 11, 6 – 21 mL  Median, IQR | 6 ± 8 mL | no |
| Wardlaw et al., 2017^34^ | Cohort | Stroke | - | 190 (190) | 65.3 (11.3) | 112 (59) | 1 | mL, FLAIR, T1. 1.5T | - | 22.0 ± 24.8 mL  Range: 0 – 121.48 mL  Median, IQR 12.56,  4.00 – 32.70 mL | 1.27 ± 8.46 mL  Range: -31.97 – 29.11 mL  Median, IQR  1.42, 22.33 – 4. 58 mL | yes |
| Whitman et al., 2001^36^ | Cohort | Healthy/CD | - | 70 (81) | 79 (4) | - | 4 | cm^2^, T2, 1.5T | - | 3.1 ± 2.5 mL | 1.1 ± 1.8 mL | no |
| Xiong et al., 2014^39^ | Case-control | Stroke | Statin use before stroke | 47 (47) with WMH volumes | 73.59 (7.82) | 28 (54.9) | 2 | cm3, FLAIR, 1.5T | Reduced progression: prestroke statin use | 25.6 ± 11.1 mL | 1.54 ± 4.52 mL | no |
| Xiong et al., 2014^39^ | Case-control | Stroke | No statin use before stroke | 28 (28) with WMH volumes | 76.87 (7.87) | 16 (53.3) | 2 | cm3, FLAIR, 1.5T | Reduced progression: prestroke statin use | 26.7 ± 19.2 mL | 5.01 ± 6 mL | no |
| Yoon et al., 2017^40^ | Non-randomized trial | Other VRF | Non-antiplatelet group | 39 (39) | 60.9 (8.7) | 11 (28.2) | 5.2^b^ | mm3, T1, T2, FLAIR, 1.5T | - | 1602.45, 619.52 – 2375.58 mm^3^  Median, IQR | 0.77 ± 1.00 mL  Median, IQR^c^: 619.19, 194.46 - 1504.42 mm^3^ | no |
| Yoon et al., 2017^40^ | Non-randomized trial | Other VRF | Antiplatelet group | 54 (54) | 60.5 (6.5) | 22 (40.7) | 5.2^b^ | mm3, T1, T2, FLAIR, 1.5T | - | 2169.64, 1165.09 – 3874.43 mm^3^  median, IQR | 1.19 ± 1.57 mL  Median, IQR^c^: 808.53, 356.58 – 2417.02 mm^3^ | no |

^a^at baseline; ^b^converted from days; ^c^original data; ^d^converted from months; ^e^converted from weeks; ^f^ additional data provided by author.

AD: Alzheimer’s disease; DLB: Lewy Body dementia; FLAIR: Fluid-attenuated inversion recovery; Healthy/CD: Healthy/community-dwelling; IQR: interquartile range; Other VRF: Other vascular risk factors; PDD: Parkinson’s disease dementia; SEM: Standard error of the mean; WMH: White matter hyperintensities

**eTable 4: Summary of included papers – WMH volume as %ICV**

| **Author, year** | **Study design** | **Population** | **Sub-group** | **N (^a^)** | **Mean age (SD)^a^** | **Sex, male**  **(%)^a^** | **Mean time between scans, years** | **WMH measure, sequence, scanner** | **Predictors**  **of WMH change** | **Baseline WMH volume, mean** **± SD** | **WMH change,**  **mean ± SD** | **WMH regression mentioned by authors** |
| --- | --- | --- | --- | --- | --- | --- | --- | --- | --- | --- | --- | --- |
| De Bresser et al., 2010^6^ | Case-control | Other VRF | DM | 55 (55) | 65.9 (5.4) | 26 (47.3) | 4.1 | %ICV, FLAIR, T1, 1.5T | - | 0.39 ± 0.78 %ICV | 0.14 ± 0.18 %ICV | no |
| De Bresser et al., 2010^6^ | Case-control | Healthy/CD | Controls | 28 (28) | 64.2 (4.3) | 12 (42.9) | 4.1 | %ICV, FLAIR, T1, 1.5T | - | 0.28 ± 0.42 %ICV | 0.12 ± 0.28 %ICV | no |
| Dickie et al., 2016^8^ | Cohort | Healthy/CD | - | 351 (351) | 72.71 (0.72) | 202 (57.5) | 3 | %ICV, FLAIR, T1, T2, T2*, 1.5T | Progression: Age | 0.76 ± 0.71 %ICV | 0.26 ± 0.28 %ICV | no |
| Moscufo et al., 2012^20^ | Cohort | Healthy/CD | - | 77 (99) | 82 (4) | 31 (60) | 1.9 | %ICV, FLAIR, 3T | - | 1.0 ± 0.9 % ICV  Range:  0.02 - 4.2 | 0.4 ± 0.5 %ICV  Range:  -0.2, 2.1 | no |
| White et al., 2019^35^ | Randomized trial | Other VRF | Standard hypertension treatment | 83 (100) | 80.3 (3.8) | 49 (49) | 3^d^ | %ICV, T1, FLAIR, 1.5T | Reduced progression: Intensive lowering of ambulatory hypertension | 0.93,  0.59 – 1.80 %ICV  Median, IQR | 0.5 ± 0.7 %ICV | no |
| White et al., 2019^35^ | Randomized trial | Other VRF | Intensive | 78 (99) | 80.9 (4.4) | 42 (42.4) | 3^d^ | %ICV, T1, FLAIR, 1.5T | Reduced progression: Intensive lowering of ambulatory hypertension | 1.17, 0.61 – 1.95 %ICV  Median, IQR | 0.29 ± 0.39 %ICV | no |
| Wyss et al., 2019^37^ | Cohort | Stroke | - | 208 (317) | 66.4 (11.3) | 129 (62) | 2 | Ratio (WMH/ ICV), T1, FLAIR, 1T and 1.5T | - | 0.0091 ± 0.0095 ratio^f^ | 0.12 ± 0.37 %ICV  Mean ± SD:  0.0012 ± 0.0037 ratio^f^ | no |
| Xia et al.,2020^38^ | Cohort | Healthy/CD | - | 191 (191) | 68.1 (63-72.6; median IQR) | 83 (43.5) | 6.7 | %ICV, FLAIR, 1.5T and 3T | Progression: High baseline WMH volume | 0.29, 0.15 – 0.52 %ICV  Median, IQR | 0.25 ± 0.28 %ICV  Median, IQR: 0.24, 0.07 – 0.45 | no |

^a^at baseline; ^b^converted from days; ^c^original data; ^d^converted from months; ^e^converted from weeks; ^f^ multiplied by 100 to create %ICV.

DM: Diabetes mellitus; FLAIR: Fluid-attenuated inversion recovery; Healthy/CD: Healthy/community-dwelling; ICV: Intracranial volume; Other VRF: Other vascular risk factors; WMH: White matter hyperintensities

**eTable 5: Summary of included papers – WMH volume as mL per year**

| **Author, year** | **Study design** | **Population** | **Sub-group** | **N (^a^)** | **Mean age (SD)^a^** | **Sex, male (%)^a^** | **Mean time between scans, years** | **WMH measure, sequence, scanner** | **Predictors**  **of WMH change** | **Baseline WMH volume, mean** **± SD** | **WMH change,**  **mean ± SD** | **WMH regression mentioned by authors** |
| --- | --- | --- | --- | --- | --- | --- | --- | --- | --- | --- | --- | --- |
| Akoudad et al., 2014^1^ | Cohort | Healthy/CD | - | 776 (831) | 68.3 (6.2) | 393 (48.9) | 3.4 | mL/year, T1, FLAIR, 1.5T | Progression: Incident microbleeds in cortico-subcortical regions | 3.7, 2.2 – 7.1 mL  Median, IQR | 0.25 ± 0.37 mL/year  Median, IQR^c^:  0.18, 0.03 – 0.53 | no |
| Carmichael et al., 2010^4^ | Case-control | Healthy/CD | Cognitively normal | 224 (224) | 76 (4.8) | 112 (52) | 1 | cm^3^/year, T1, T2, proton density, 1.5T | - | 0.51 ± 1.1 mL | 0.082 ± 0.92 mL/year | no |
| Carmichael et al., 2010^4^ | Case-control | Cognitive | MCI | 391 (391) | 75 (7.5) | 240 (65) | 1 | cm^3^/year, T1, T2, proton density, 1.5T | - | 0.66 ± 1.2 mL | 0.24 ± 1.2 mL/year | no |
| Carmichael et al., 2010^4^ | Case-control | Cognitive | AD | 189 (189) | 76 (7.5) | 94 (53) | 1 | cm^3^/year, T1, T2, proton density, 1.5T | - | 1.1. ± 2 mL | 0.24 ± 1.5 mL/year | no |
| Chen et al., 2006^5^ | Cohort | Stroke | CAA | 26  (26) | 69.1 (6.5) | 13 (50) | 1.1 (median) | mL/year, T2, FLAIR, 1.5T | Progression: Baseline WMH volume | 4.1,  2.1 – 25.1 mL  Median, IQR | 1.13 ± 2.12 mL/year  Median, IQR^c^:  0.5, 0.1 – 2.8 | no |
| Maillard et al., 2013^17^ | Case-control | Healthy/CD | Cognitively normal | 72  (72) | 74.2 (6.6) | 20 (27.8) | 4.1 | mL/year, FLAIR, 1.5T | Progression: Baseline FA, and FLAIR signal intensity | 6.6. ± 9.5 mL | 0.41 ± 1.4 mL/year | no |
| Maillard et al., 2013^17^ | Case-control | Cognitive | MCI | 34  (34) | 74.7 (8.6) | 15 (44.1) | 2.9 | mL/year, FLAIR, 1.5T | Progression: Baseline FA, and FLAIR signal intensity | 9.4 ± 10.0 mL | 0.47 ± 1.56 mL/year | no |
| Maillard et al., 2013^17^ | Case-control | Cognitive | AD | 13  (13) | 76.4 (8.7) | 6 (46.1) | 3.3 | mL/year, FLAIR, 1.5T | Progression: Baseline FA, and FLAIR signal intensity | 9.0 ± 8.8 mL | 1.22 ± 1.41 mL/year | no |
| Sam et al., 2016^25^ | Cohort | Stroke | - | 45  (45) | 74 (9.4) | 25 (55.6) | 1 | mL/year, FLAIR, proton density, T2, 3T | - | 32 ± 25 mL | 0.66 ± 0.60 mL/year | Yes; dismissed as error |
| Scharf et al., 2019^41^ | Cohort | Healthy/CD | 60 – 69 years of age | 247 (247) | 65.3 (2.6) | 126 (51) | 2.6 | cm^3^/year, T1, FLAIR, 3T | Progression: Age, baseline WMH, baseline hypertension, midlife hypertension, baseline fasting glucose in males | 0.61 ± 0.76 %ICV | 0.54 ± 1.27 mL/year | no; some individual trajectories do show regression |
| Scharf et al., 2019^41^ | Cohort | Healthy/CD | 70 – 79 years of age | 186 (186) | 74.9 (3) | 97 (52) | 3.1 | cm^3^/year, T1, FLAIR, 3T | Progression: Age, baseline WMH, baseline hypertension, midlife hypertension, baseline fasting glucose in males | 1.08 ± 0.85 %ICV | 1.04 ± 1.93 mL/year | no; some individual trajectories do show regression |
| Scharf et al., 2019^41^ | Cohort | Healthy/CD | ≥80 years of age | 121 (121) | 84.8 (3.5) | 76 (63) | 2.8 | cm^3^/year, T1, FLAIR, 3T | Progression: Age, baseline WMH, baseline hypertension, midlife hypertension, baseline fasting glucose in males | 1.81 ± 1.24 %ICV | 1.6 ± 2.4 mL/year | no; some individual trajectories do show regression |
| Van Dalen et al., 2017^30^ | Randomized trial | Other VRF | Standard care | 62  (63) | 77.1 (2.4) | 29 (47) | 2.8^d^ | mL/year, FLAIR, 3T | Progression: Baseline WMH volume, systolic BP, diastolic BP, mean arterial pressure, BMI | 5.7,  3.3 – 11.1 mL  Median, IQR | 0.7 ± 0.59 mL/year | no |
| Van Dalen et al., 2017^30^ | Randomized trial | Other VRF | Vascular care | 64  (72) | 77.3 (2.6) | 30 (47) | 2.8^d^ | mL/year, FLAIR, 3T | Progression: Baseline WMH volume, systolic PB, diastolic BP, mean arterial pressure, BMI | 6.3,  3.5 – 10.9 mL  Median, IQR | 0.73 ± 0.84 mL/year | no |

^a^at baseline; ^b^converted from days; ^c^original data; ^d^converted from months.

AD: Alzheimer’s disease; BMI: Body mass index; BP: Blood pressure; CAA: Cerebral amyloid angiopathy; FA: Fractional anisotropy; FLAIR: Fluid-attenuated inversion recovery; Healthy/CD: Healthy/community-dwelling; IQR: interquartile range; MCI: Mild cognitive impairment; Other VRF: Other vascular risk factors; WMH: White matter hyperintensities

**eTable 6: Summary of relevant papers without useable WMH change data**

| **Study** | **Study design** | **Population** | **Sub-group** | **N (^a^)** | **Mean age (SD)**^a^ | **Mean time between scans, years** | **WMH measure, sequence, scanner** | **Predictors of WMH change** | **WMH change, mean ± SD** | **WMH regression mentioned by authors** |
| --- | --- | --- | --- | --- | --- | --- | --- | --- | --- | --- |
| Bahrani et al., 2021^42^ | Cohort | Healthy/CD and cognitive | - | Total 79 (79)  Healthy/CD 39  Cognitive 40 | 74.01 (7.6) | 1 | T1, FLAIR, 3T | - | - | yes; 19% of participants shows overall regression. 31% of participants had areas of regression and progression |
| Cho et al., 2015^43^ | Cohort | Stroke | - | 100 (100) | 67.5 (11.8) | 2.3 ^c^ | cc,  T2, FLAIR, - | Progression: age, male sex, large vessel disease, poor renal function  Regression: no significant factors | Patients with progression: 3.32 (0.92 - 4.05) cc, median (IQR)  Patients with regression: -1.87 cc (-4.61 to -0.73) | yes; 21.5% of patients had WMH regression |
| Dadar et al., 2020^44^ | Cohort | Healthy/CD | - | 207 (207) | 75.7 (5.7) | 1 | mm^3^, T1, T2/PD, FLAIR, - | Progression: higher baseline WMH volume and lower amyloid-beta1-42 level | - | no; figures show WMH regression |
| Dadar et al., 2020^44^ | Cohort | Cognitive | MCI | 396 (396) | 73.5 (7.4) | 1 | mm^3^, T1, T2/PD, FLAIR, - | Progression: higher baseline WMH volume and lower amyloid-beta1-42 level | - | no; figures show WMH regression |
| Dadar et al., 2020^44^ | Cohort | Cognitive | AD | 117 (117) | 74.5 (7.8) | 1 | mm^3^, T1, T2/PD, FLAIR, - | Progression: higher baseline WMH volume and lower amyloid-beta1-42 level | - | no; figures show WMH regression |
| Debette et al., 2011^45^ | Cohort | Healthy/CD | - | 1352 or 1493^d^ | 61 (9) | 6.3 | cm^3^/year, T1, double echo proton density, T2, 1 or 1.5T | Progression: Hypertension and smoking in midlife | 0.22 ± 0.44 mL/year | no |
| De Groot et al., 2013^46^ | Cohort | Healthy/CD | - | 689 (722) | 66.9 (5.0) | 3.5 | mL, T1, FLAIR, DTI. 1.5T | Progression: Lower FA, higher MD and relatively higher normalized FLAIR intensity at baseline | Median (IQR) decrease “lost” 0.8 mL (0.5-1.3)  Median (IQR) increase “new” 1.4mL (0.8-2.8)  Median (IQR) increase “de novo” 0.2mL (0.1-0.3) | yes; net decrease of WMH volume in 19% of participants explained as misclassification of tissues and measurement error |
| Erdélyi-Bótor et al., 2015^47^ | Cohort | Healthy/CD | - | 17 (17) | 47 (11.2) | 3^c^ | mL, T1, T2, FLAIR, 3T | Disappearance of WMH more common in a migraine with low attack frequency | 91 WMH (23.3%) decreased in size. All participants had at least 1 WMH that decreased.  22 WMH disappeared | yes; WMH that decreased in size and some disappeared |
| Findlay et al.,2019^48^ | Cohort | Other VRF | - | 34 (97) | 58 (50.5 – 66.5), median (IQR) | 1.05^c^ (median) | mL, T1, T2, FLAIR, 3T | - | Patients with continued hemodialysis had WMH volume increase: mean 2.96 mL to 3.36 mL  Patient with transplant had no significant WMH volume increase: mean 1.02 mL to 1.17 mL | no |
| Gu et al., 2019^49^ | Cohort | Healthy/CD and Cognitive | C-reactive protein | 508 (680)  45 demented | 79.92 (5.68) | 4.57^c^ | Log_10_,  T1, FLAIR, T2, 1.5T | Progression: associated with Log_10_ACT | 0.62 ± 0.4 (log_10_) | no |
| Gu et al., 2019^49^ | Cohort | Healthy/CD and Cognitive | Interleukin-6 (IL6) | 435 (680)  45 demented | 79.73 (5.48) | 4.52^c^ | Log_10_,  T1, FLAIR, T2, 1.5T | Progression: associated with Log_10_ACT | 0.61 ± 0.41 (log_10_) | no |
| Gu et al., 2019^49^ | Cohort | Healthy/CD and Cognitive | Alpha 1-antichymotrypsin (ACT) | 435 (680)  45 demented | 79.99 (5.68) | 4.52 ^c^ | Log_10_,  T1, FLAIR, T2, 1.5T | Progression: associated with Log_10_ACT | 0.61 ± 0.4 (log_10_) | no |
| Jochemsen et al., 2012^50^ | Cohort | Other VRF | - | 682 (682 for analysis) | 57.5 (9.5) | 3.9^c^ | %ICV. T1, T2, FLAIR and IR, 1.5T | DWMH progression: Higher ACE activity possibly associated | Total WMH:0.02% ±0.32%  PVWMH: 0.03 ± 0.15%  DWMH: -0.01% ±0.22% | no; mean DWMH volume change suggest very minor regression |
| Kim et al., 2020^51^ | Cohort | Cognitive | WMH progression group | 70 (70) | 72.1 (6.5) | 3^c^ | mL, T1, FLAIR, T2, 3T | - | Increase: 0.29 mL per year. | yes |
| Kim et al., 2020^51^ | Cohort | Cognitive | WMH regression group | 17 (17) | 71.8 (8.7) | 3^c^ | mL, T1, FLAIR, T2, 3T | - | Decrease: 0.06 mL per year | yes |
| Markus et al., 2021^52^ | Randomized trial | Stroke | Standard treatment | 39 (56) | 69.6 (9.4) | 2 | %whole brain volume, FLAIR, 3T | Less progression: More intensive BP lowering | 0.6 ± 0.7%brain | no |
| Markus et al., 2021^52^ | Randomized trial | Stroke | Intensive treatment | 42 (55) | 68.1 (8.7) | 2 | %whole brain volume, FLAIR, 3T | Less progression: More intensive BP lowering | 0.4 ± 0.9 %brain | no |
| Mok et al., 2009^53^ | Randomized trial | Other VRF | Intervention (simvastatin) | 106 (106) | 60.1 (9.5) | 2^c^ | cm^3^, visual, Turbo spin echo T2, 1.5T | Progression: Baseline WMH volume  Delay of progression (with severe WMH at baseline): simvastatin treatment | WMH volume increase: 0.0 (0.3) cm^3^, median (IQR)  Change in severe WMH group: 1.9 (2.5) cm^3^, median (IQR) | no |
| Mok et al., 2009^53^ | Randomized trial | Other VRF | Placebo | 102 (102) | 61.7 (9.0) | 2^c^ | cm^3^, visual, Turbo spin echo T2, 1.5T | Progression (with severe WMH at baseline): Baseline WMH volume  Delay of progression: simvastatin treatment | WMH volume increase: 0.0 (0.0) cm^3^, median (IQR)  Change in severe WMH group: 3.0 (3.9) cm^3^, median (IQR) | no |
| Nasrallah et al., 2019^54^ | Randomized trial | Other VRF | Intensive treatment  (SBP <120 mmHg) | 249 (355) Baseline 355  Follow-up 249 included for analysis | 67.7 (8.0) | 3.97^c^ | Estimated and asinh volume, cm^3^,T1, T2, FLAIR, 3T | - | Estimated change: 0.92 (0.69-1.14), median (IQR)  Estimated annualized change: 0.23 (0.17-0.29), median (IQR) | no |
| Nasrallah et al., 2019^54^ | Randomized trial | Other VRF | Standard treatment  (SBP <140 mmHg) | 200 (315) | 66.9 (8.5) | 3.97^c^ | Estimated and asinh volume, T1, T2, FLAIR. 3T | - | Estimated change 1.45 (1.21-1.70) median (IQR)  Estimated annualized change 0.37 (0.30-0.43), median (IQR) | no |
| Nylander et al., 2018^55^ | Cohort | Healthy/CD | - | 252 (406) | 75 | 5^c^ | Visual, mL, T1, T2, T2*, 1.5T | - | Increase of 1.8 mL | yes; Dismissed as measurement error |
| Peng et al., 2014^56^ | Cohort | Other VRF | - | 250 (25) | 84.42 (2.52) | 4^c^ | mL, FLAIR, T1, 3T | Progression: High (>160 mm Hg) and low SBP (<140 mm Hg) levels | WMH % change: 29.96% ± 15.18%,  WMH/TIV ratio change:  64.01% ± 28.89% | no |
| Promjunyakul et al., 2018^57^ | Cohort | Healthy/CD | - | 52 (120) | 82.8 (7.6) | 1.4^b^ | mL, T1, FLAIR, 3T | Progression:  Low baseline CBF and FA, and high MD, AD and RD | PVWMH:  3.7 ± 6 mL  DWMH:  0.4 ± 1 mL | no |
| Silbert et al., 2012^58^ | Cohort | Healthy/CD | Converters to MCI | 134 (181) | 84.5 (5.9) | 10.1^c^ | Natural log transformed WMH, T1, T2, 1.5T | - | Annual change in log transformed WMH 0.062 (SE 0.015)  Annual change rate before onset to MCI 6.5%  Additional change log transformed WMH 0.032 (SE 0.016)  Additional rate of annual change after conversion 3.3%. | no |
| Silbert et al., 2012^58^ | Cohort | Healthy/CD | Non-converters to MCI | 47 (181) | 82.1 (8.1) | 6.9^c^ | Natural log transformed WMH,. T1, T2, 1.5T | - | Annual rate of %WMH change increased after MCI onset | no |
| Van den Heuvel et al., 2006^59^ | Randomized trial (substudy) | Other VRF | - | 554 (554) | 75 (3.2) | 3^c^ | mL, FSE and FLAIR. 1.5T | - | - | no |
| Van Overbeek et al., 2016^60^ | Cohort | Stroke | - | 132 (149)  Included in analysis: 127 | 62.5 (11.7) | 2^c^ | Visual, T2 and FLAIR, 1.5T and 3T. | PWMH progression: Plasma t-PA activity | WMH progression in 61 (48%) patients | no |
| Wang et al., 2012^61^ | Cohort | Stroke | - | 46 (46) | 68 (11) | 3.25^c^ | mL, T1, T2, T2*, FLAIR, 1.5T | - | WMH volume increased in 43 patients and decreased in 3 patients, range -1.94 to 33.16 mL (excluding stroke lesions) | yes |
| Wang et al., 2017^62^ | Randomized trial | Cognitive | RIC (remote ischemic conditioning) | 14 (18) | 65.71 (8.49) | 1^c^ | cm^3^. T1, T2, FLAIR, 3T | - | Change 2.632 cm^3^ | yes; RIC arm significantly reduced WMH volume. Controls reduced but not significantly |
| Wang et al., 2017^62^ | Randomized trial | Cognitive | Controls | 16 (18) | 60.75 (12.82) | 1^c^ | cm^3^. T1, T2, FLAIR. 3T | - | Change 0.935 cm^3^ | yes; Controls reduced but not significantly |
| Wolfson et al., 2005^63^ | Case-Control | Healthy/CD | Normal mobility | 7 (7) | 81 (1.7) | 1.7^b^ | %ICV/year, proton density, T2, 1.5T | - | WMH change: 0.02 ±0.05 %ICV/year | no |
| Wolfson et al., 2005^63^ | Case-Control | Healthy/CD | Impaired mobility | 7 (7) | 84 (3.4) | 1.7^b^ | %ICV/year PD,T2, 1.5T | - | WMH change: 0.10± 0.10 %ICV/year | no |
| Xu et al., 2018^64^ | Case - Control | Stroke | Lacunar stroke | 76 (-) | 72.45 (10.93) | 2^b^ | % of baseline volume, Fazekas, T1, T2, FLAIR, 1.5T or 3.0T | - | - | no; Tables 2, 3 and Figures 3B and D suggest WMH regression |
| Xu et al., 2018^64^ | Case - Control | Healthy/CD | Controls | 111 (-) | 68.37 (10.81) | 2^b^ | % of baseline volume, Fazekas, T1, T2, FLAIR, 1.5T or 3.0T | - | - | no; Tables 2, 3 and Figures 3B and D suggest WMH regression |
| Zeestraten et al., 2017^65^ | Cohort | Stroke | - | 99 (99) | 68.9 (10.0) | 3^c^ | WMH load (log), T1, FLAIR, T2*, 1.5T | - | Estimated mean annual change 0.082 ±0.033 WMH lesion load (log) | no |

^a^at baseline; ^b^converted from months; ^c^original data; ^d^Discrepancy in N between main body paper and supplemental data.

ACE: Angiotensin-converting-enzyme; AD: Axial diffusivity; BP: Blood pressure; CBF: Cerebral blood flow; DWMH: Deep white matter hyperintensities; FA: Fractional anisotropy; FLAIR: Fluid-attenuated inversion recovery; FSE: Fast spin echo; Healthy/CD: Healthy/community-dwelling; ICV: Intracranial volume; IQR: Interquartile range; MD: Mean diffusivity; MCI: Mild Cognitive Impairment; Other VRF: Other vascular risk factors; PVWMH: Periventricular white matter hyperintensities; RD: Radial diffusivity; SBP: Systolic blood pressure; SE: Standard error; TIA: Transient ischaemic attack; TIV: Total intracranial volume; tPA: Tissue plasminogen activator; WMH: White matter hyperintensities; WMSA: White matter signal abnormality

**eResult 1: WMH regression**

From relevant papers without useable WMH change data (Table S7) 3/21 paper do not mention regression but have results in figures or tables.^44, 50, 64^ 2/21 dismissed regression as an error^46, 55^ despite reporting regression in 19% of their healthy/community-dwelling participants.^46^ WMH regression was explicitly mentioned in 6/21 papers.^42, 43, 47, 51, 61, 62^ Local WMH regression was found in migraine patients^47^ and a net WMH regression was found in 6.5% and 21.5% of stroke patients,^43, 61^, in 19% of healthy/CD or participants with cognitive presentation^42^ and in 14 patients with SVD and MCI after 1 year of remote ischemic conditioning,^62^ and in 19.5% of MCI participants.^51^

**eResult 2: Summary of WMH changes**

Factors related to WMH progression included high baseline WMH^2, 5, 8, 9, 24, 30, 38, 41, 44, 53^ , older age^29, 32, 41, 43^, male sex^43^, (self-reported) diabetes^29^, hypertension^7, 9, 15, 30, 41, 45, 56^, apolipoprotein E genotype^11^, lower amyloid-beta1-42 level,^44^ smoking^45^, late life depression^14^, large vessel disease^43^, poor renal function^43^, middle cerebral artery resistance index ratio^16^, fibrinogen receptor complex on platelets, microvesicles, thrombogenic microvesicles^23^, microbleeds^1^, low cerebral blood flow^57^, low baseline fractional anisotropy^17, 46, 57^, high mean diffusivity^57^, high baseline FLAIR signal intensity^17^, fasting glucose in men^41^, high body mass index^30^, high angiotensin-converitng enzyme activity^50^, and an increase in circulating inflammatory biomarker alpha 1-antichymotrypsin^49^. Periventricular WMH progression was related to plasma t-PA activity^60^. Factors related to slowing down WMH progression were intensive blood pressure control^7, 35, 52^, use of mixed tocotrienols^12^, symptoms of cardiac disease^29^, pre-stroke statin use^39^, and simvastatin treatment for patients with high WMH burden at baseline^53^.

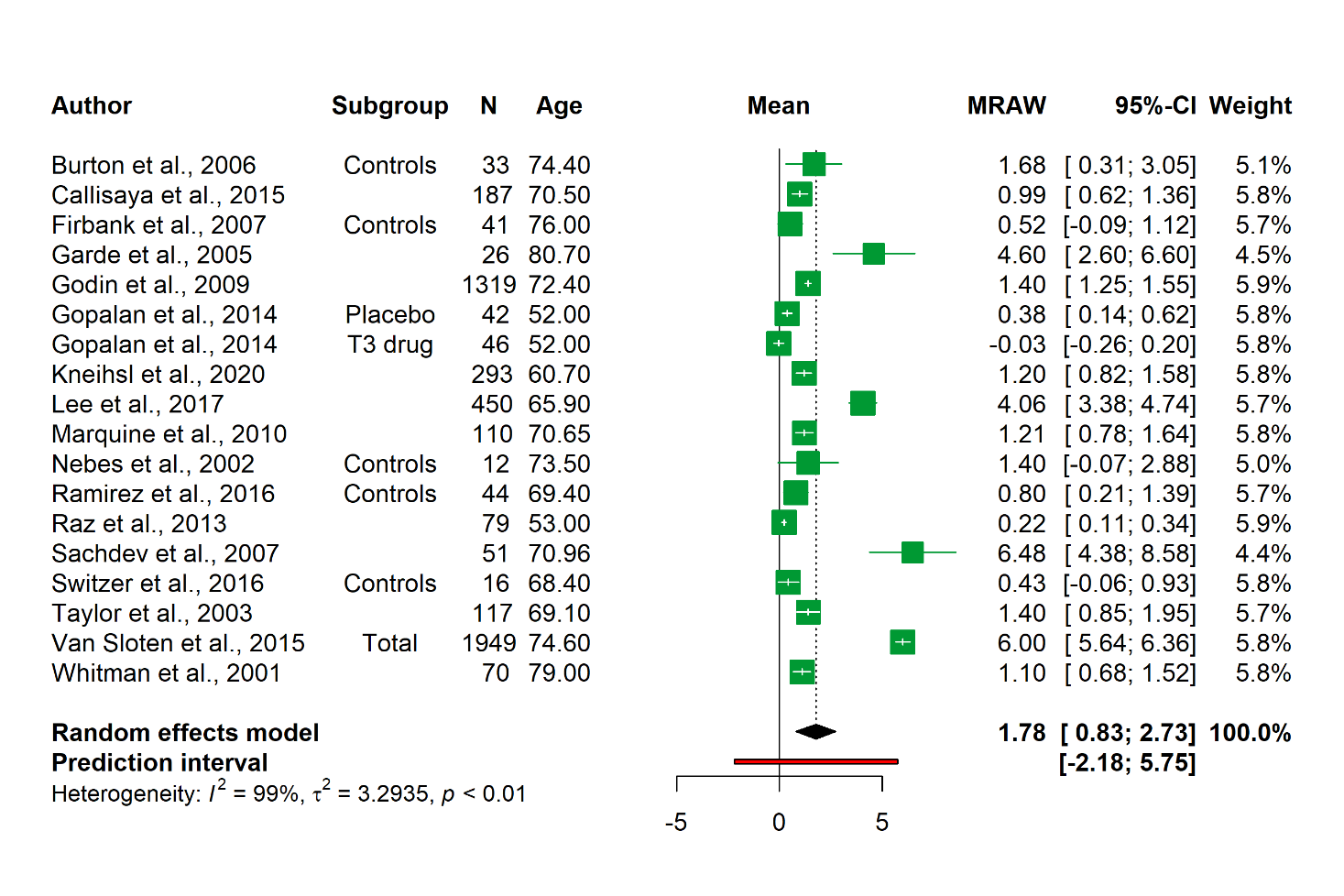
**eFigure 1. Forest plot of subgroup analysis healthy/community-dwelling population (mL)**

MRAW: raw means

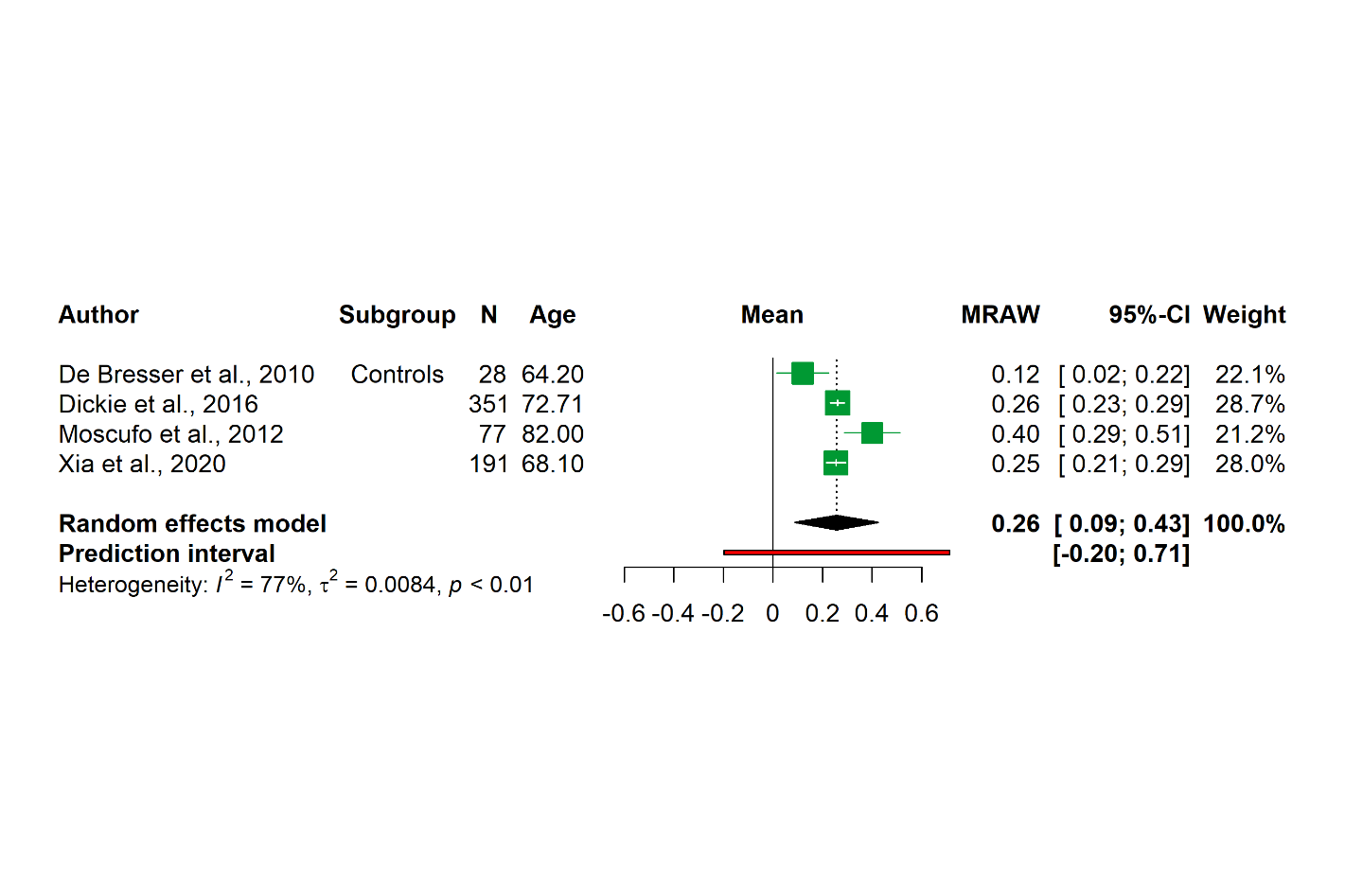
**eFigure 2. Forest plot of subgroup analysis healthy/community-dwelling population (%ICV)**

ICV: Intracranial volume; MRAW: raw means

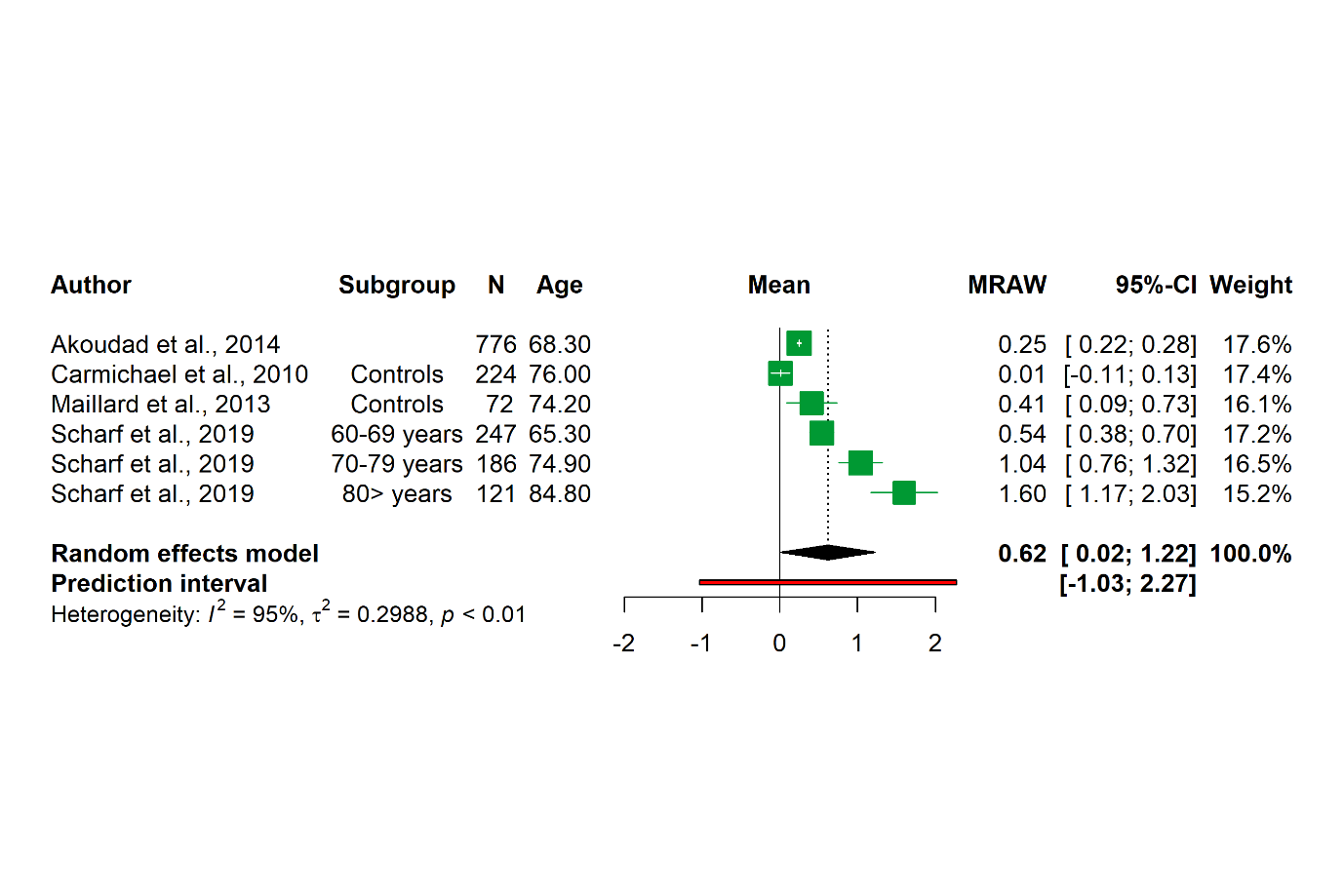
**eFigure 3. Forest plot of subgroup analysis healthy/community-dwelling population (mL/year)**

MRAW: raw means

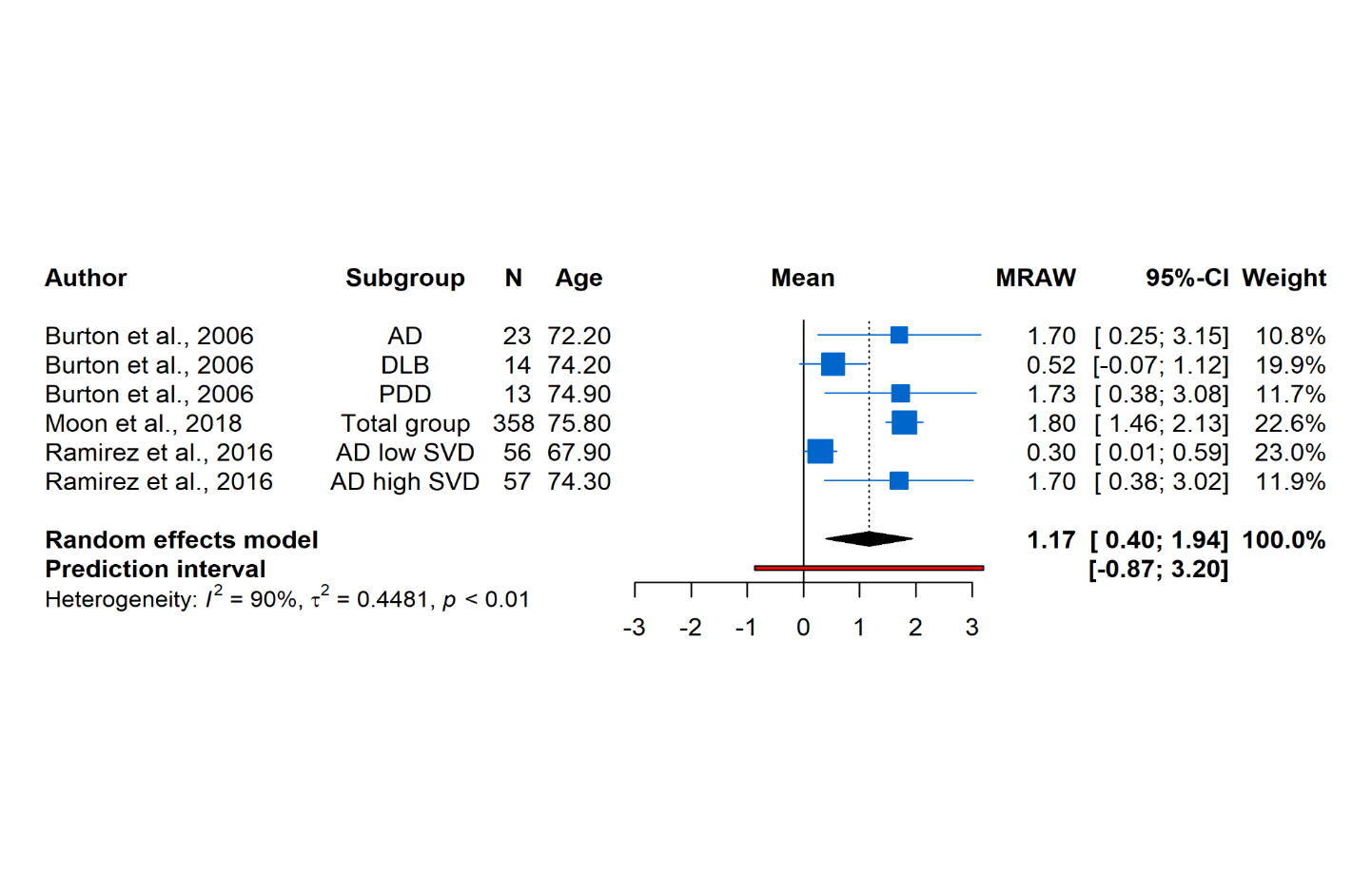
**eFigure 4. Forest plot of subgroup analysis cognitive impairment (mL)**

AD: Alzheimer’s disease; DLB: Lewy-body dementia; MRAW: raw means PDD: Parkinson’s dementia; SVD: Small vessel disease

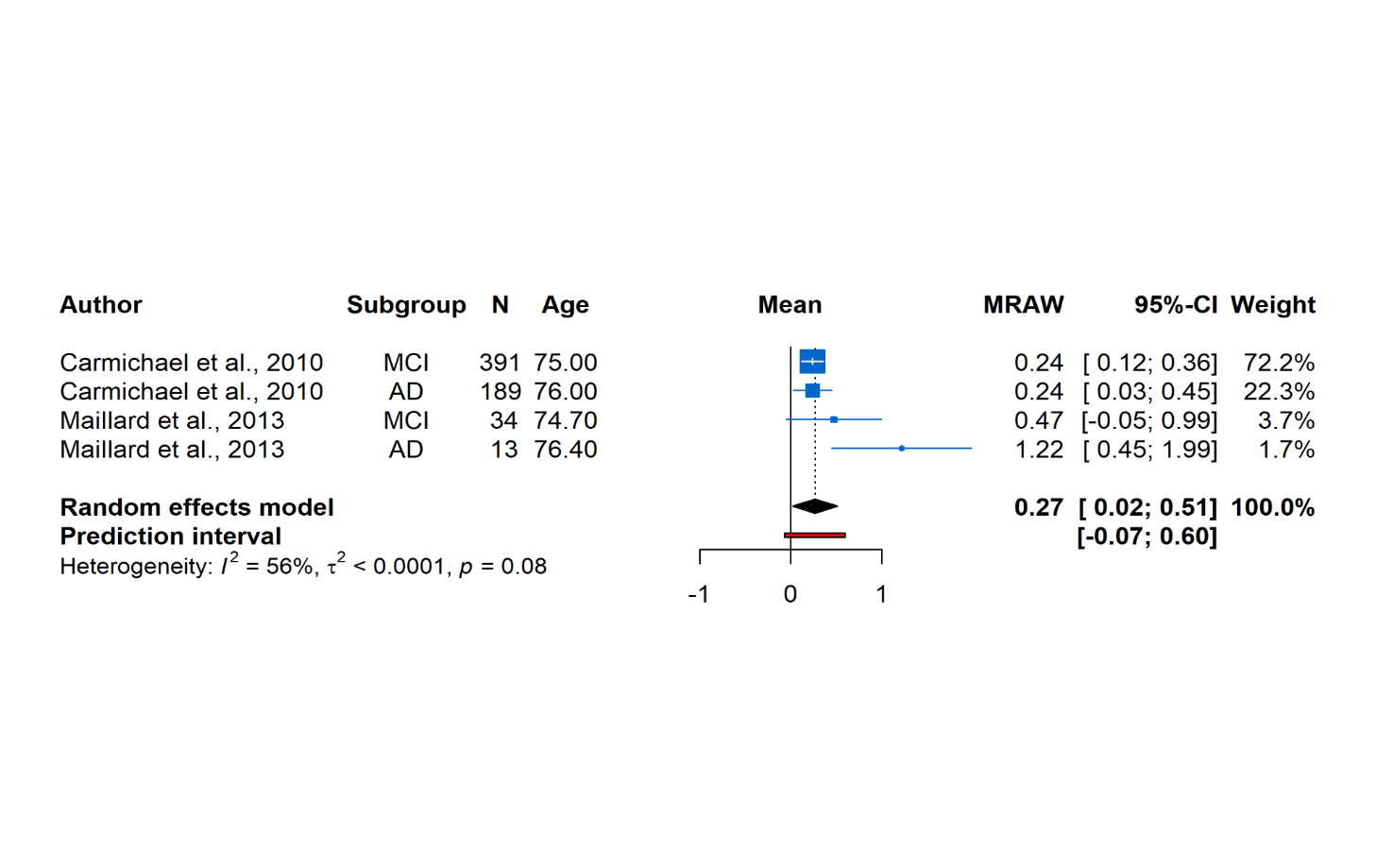
**eFigure 5. Forest plot of subgroup analysis cognitive impairment (mL/year)**

AD: Alzheimer’s disease; MCI: mild cognitive impairment; MRAW: raw means

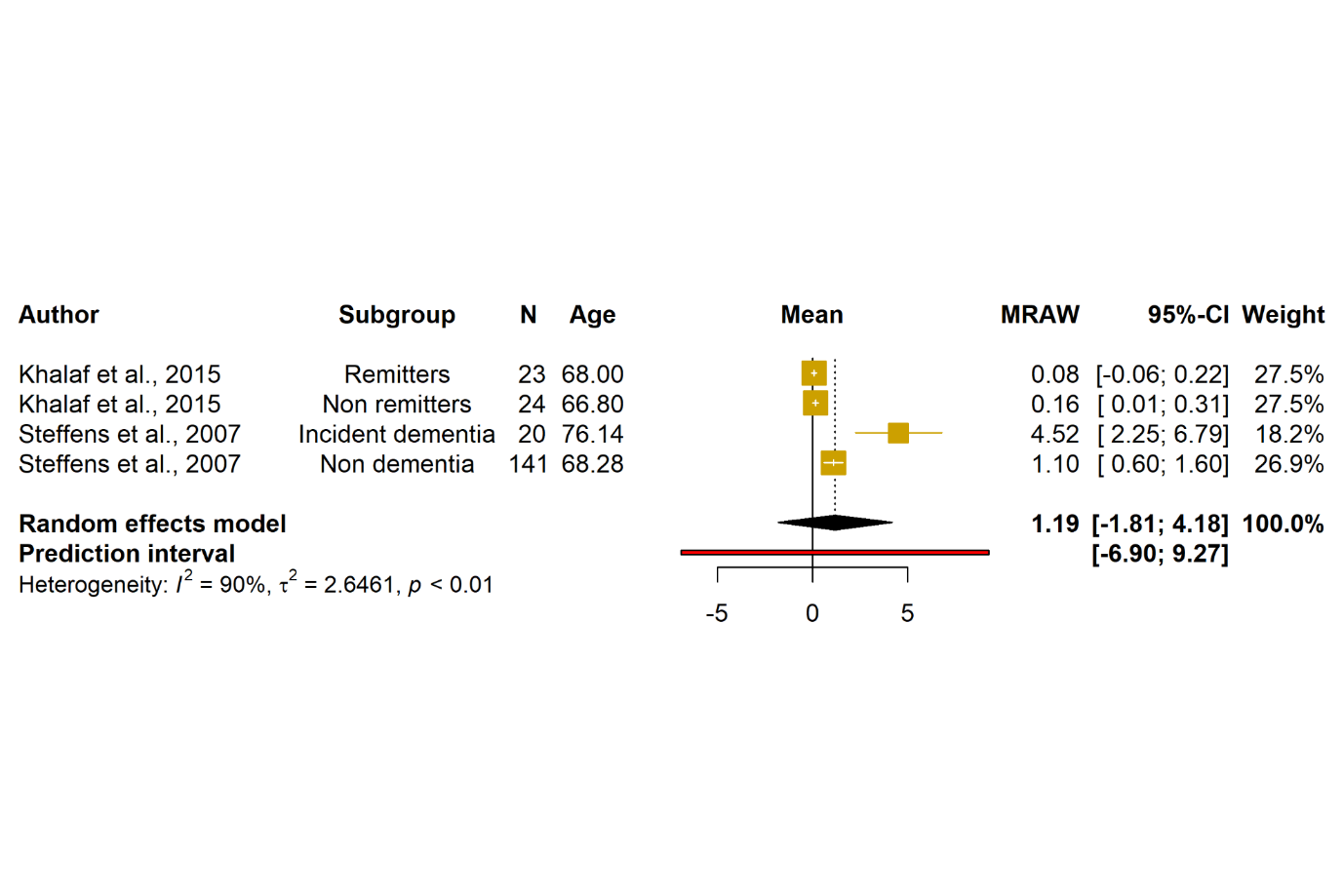
**eFigure 6. Forest plot of subgroup analysis depression (mL)**

MRAW: raw means

**eFigure 7. Forest plot of subgroup analysis other vascular risk factor (mL**
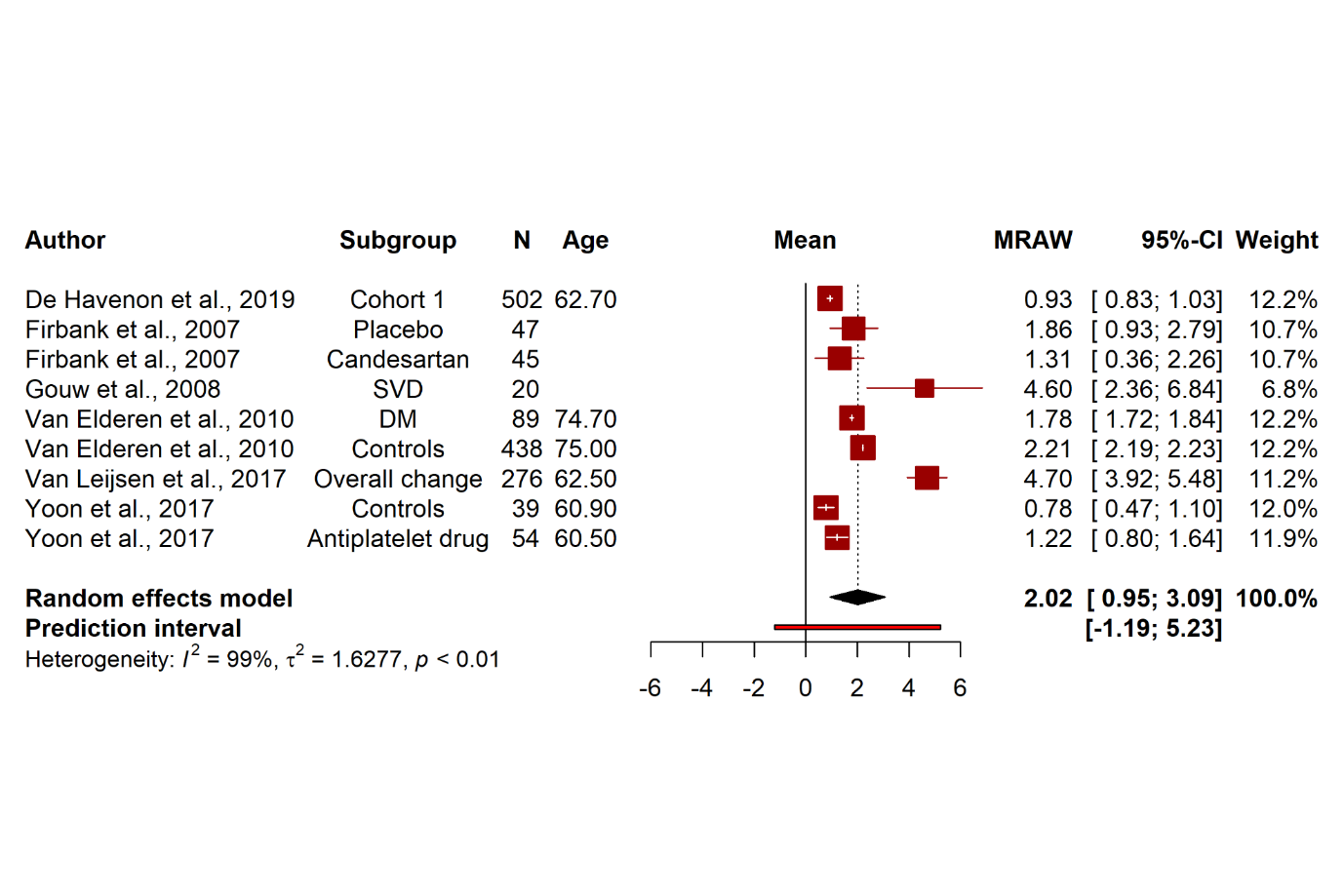
**)**

DM: Diabetes mellitus; MRAW: Raw means; SVD: Small vessel disease

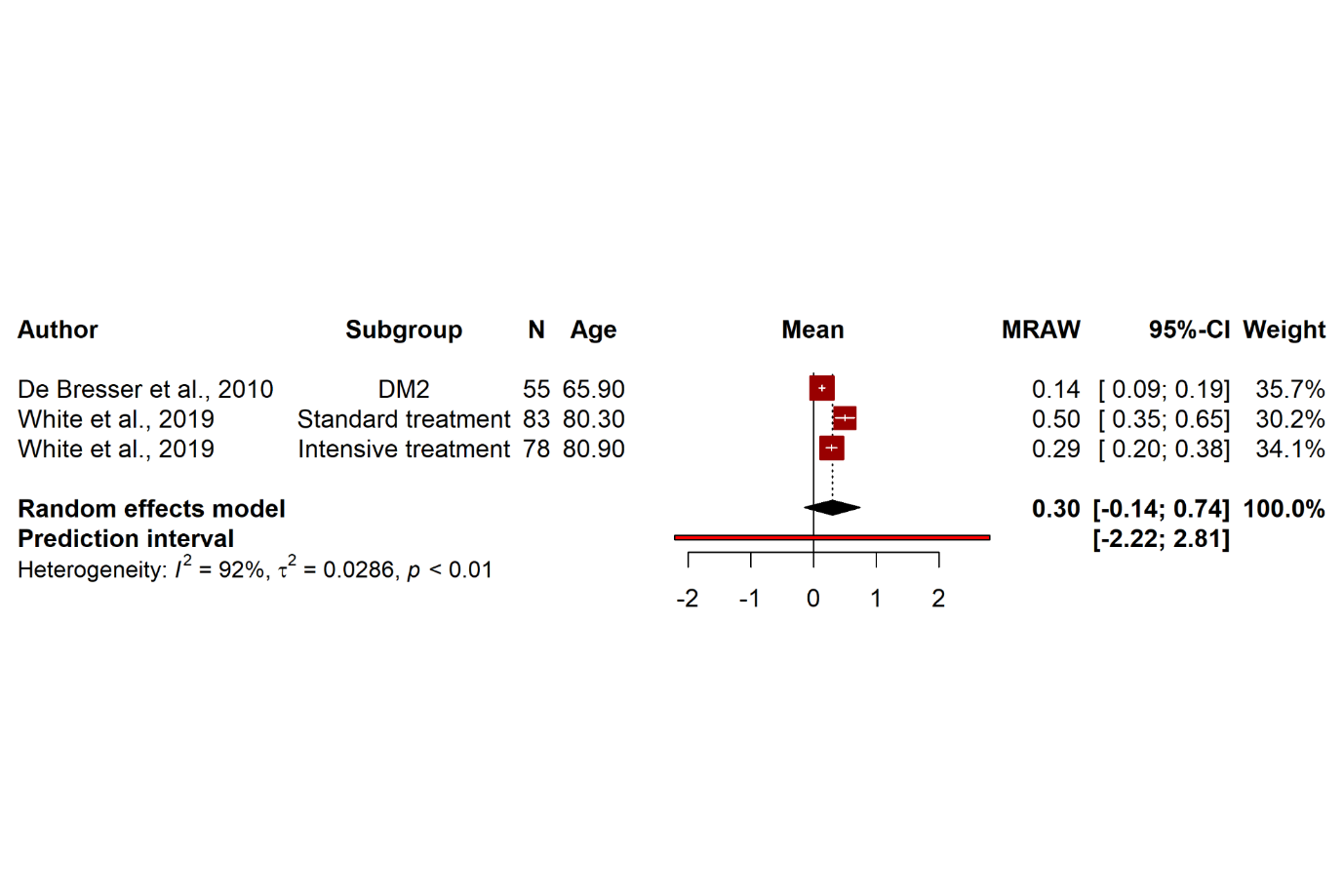
**eFigure 8. Forest plot of subgroup analysis other vascular risk factor (%ICV)**

DM2: Diabetes mellitus type 2; ICV: Intracranial volume; MRAW: raw means

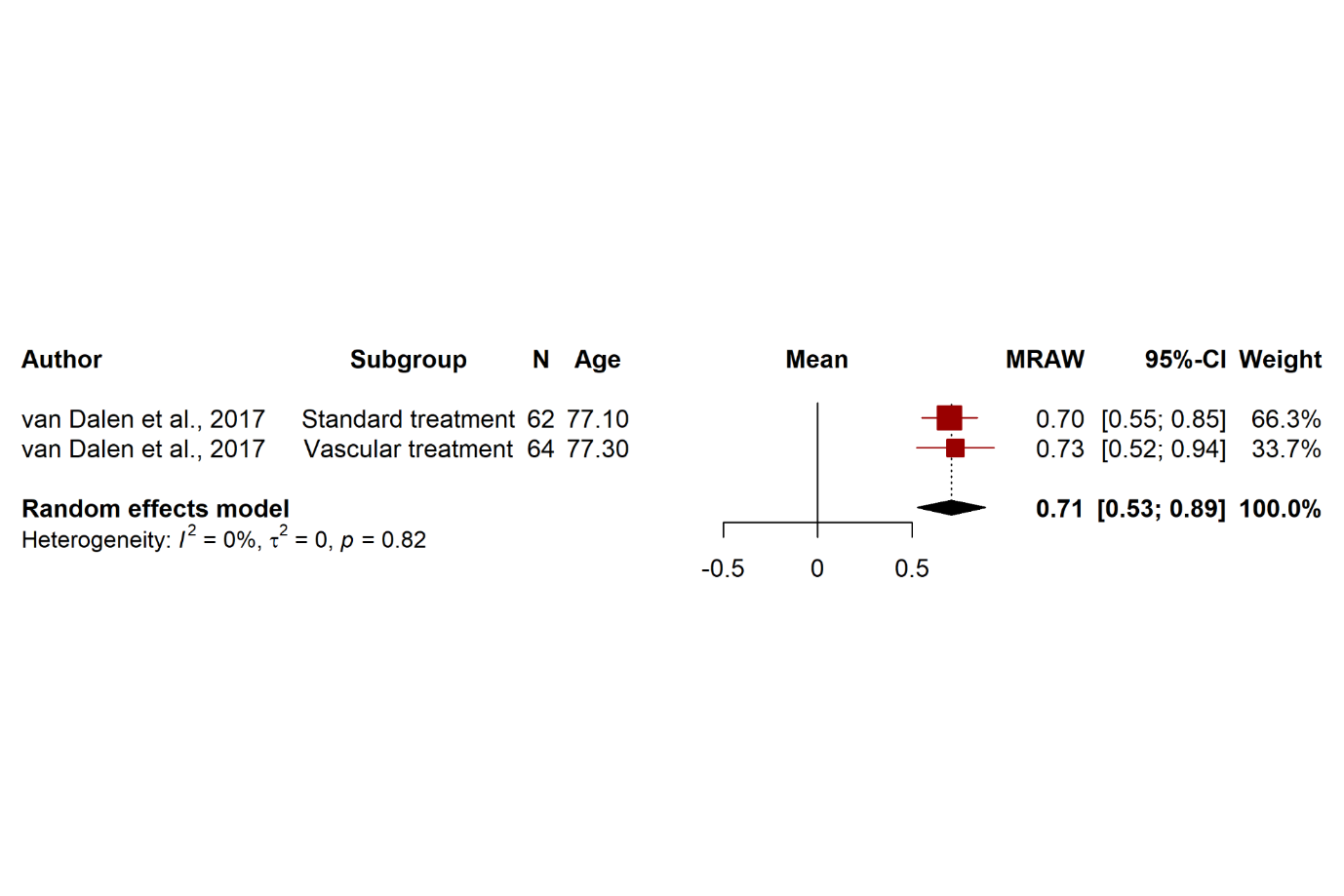
**eFigure 9. Forest plot of subgroup analysis other vascular risk factor (mL/year)**

MRAW: raw means

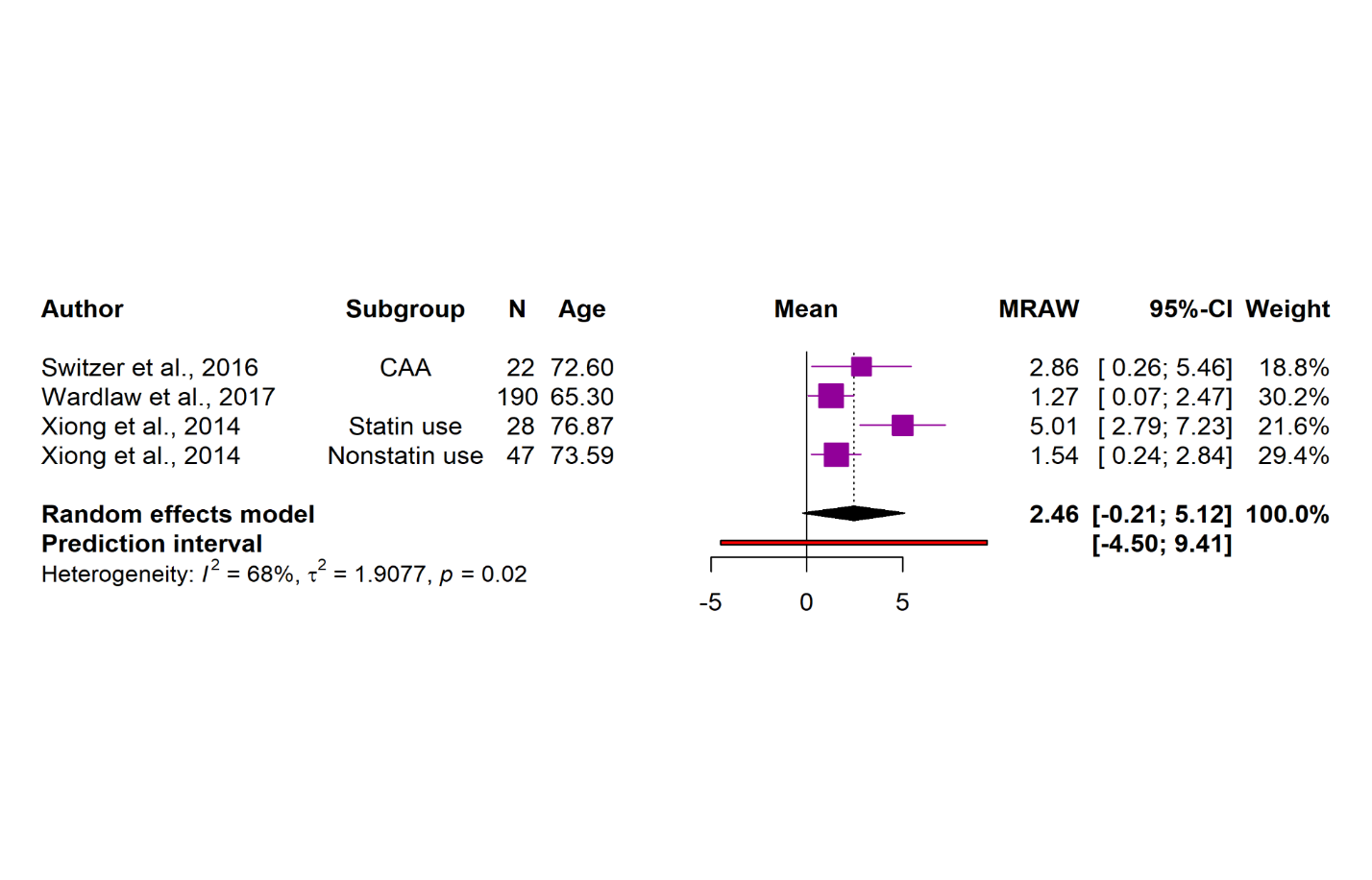
**eFigure 10. Forest plot of subgroup analysis stroke (mL)**

CAA: Cerebral amyloid angiopathy; MRAW: raw means

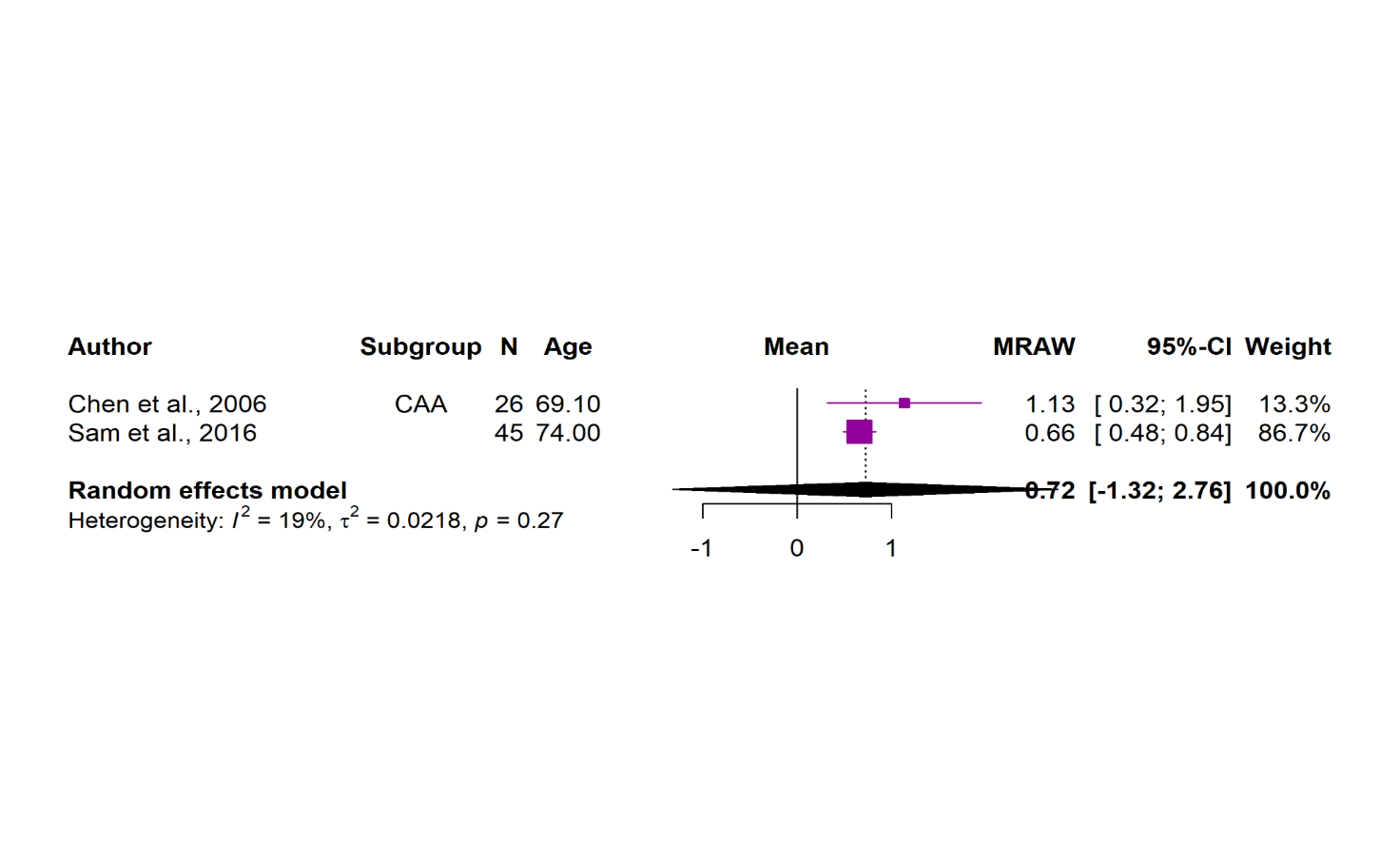
**eFigure 11. Forest plot of subgroup analysis stroke (mL/year)**

CAA: Cerebral amyloid angiopathy; MRAW: raw means

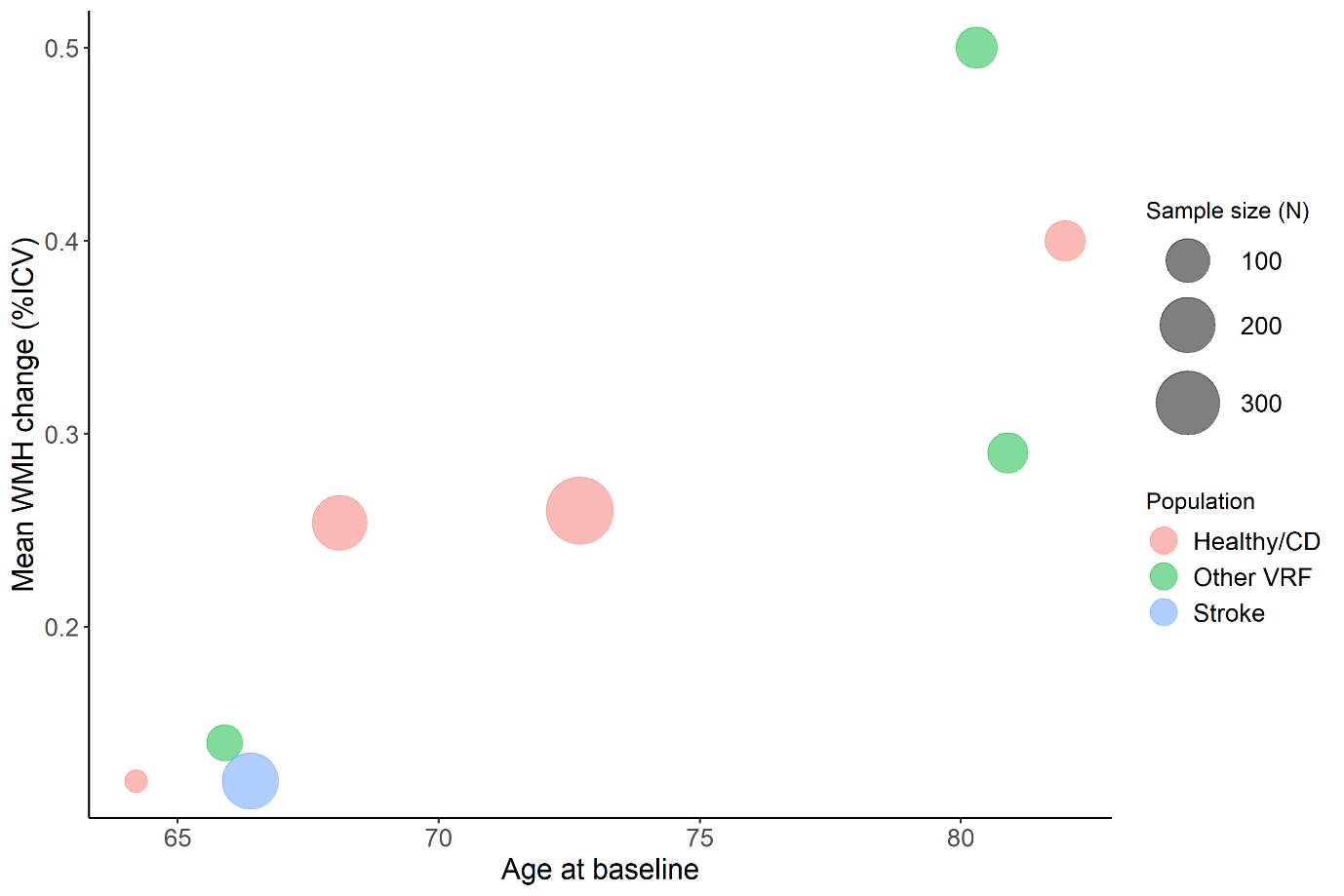
**eFigure 12. Bubble plot of mean WMH change (%ICV) related to age at time of baseline. Points in plot are scaled by sample size and colour of points refer to population group.**

e e**eFigure 13. Bubble plot of mean WMH change (mL/year) related to age at time of baseline. Points in plot are scaled by sample size and colour of points refer to population group**

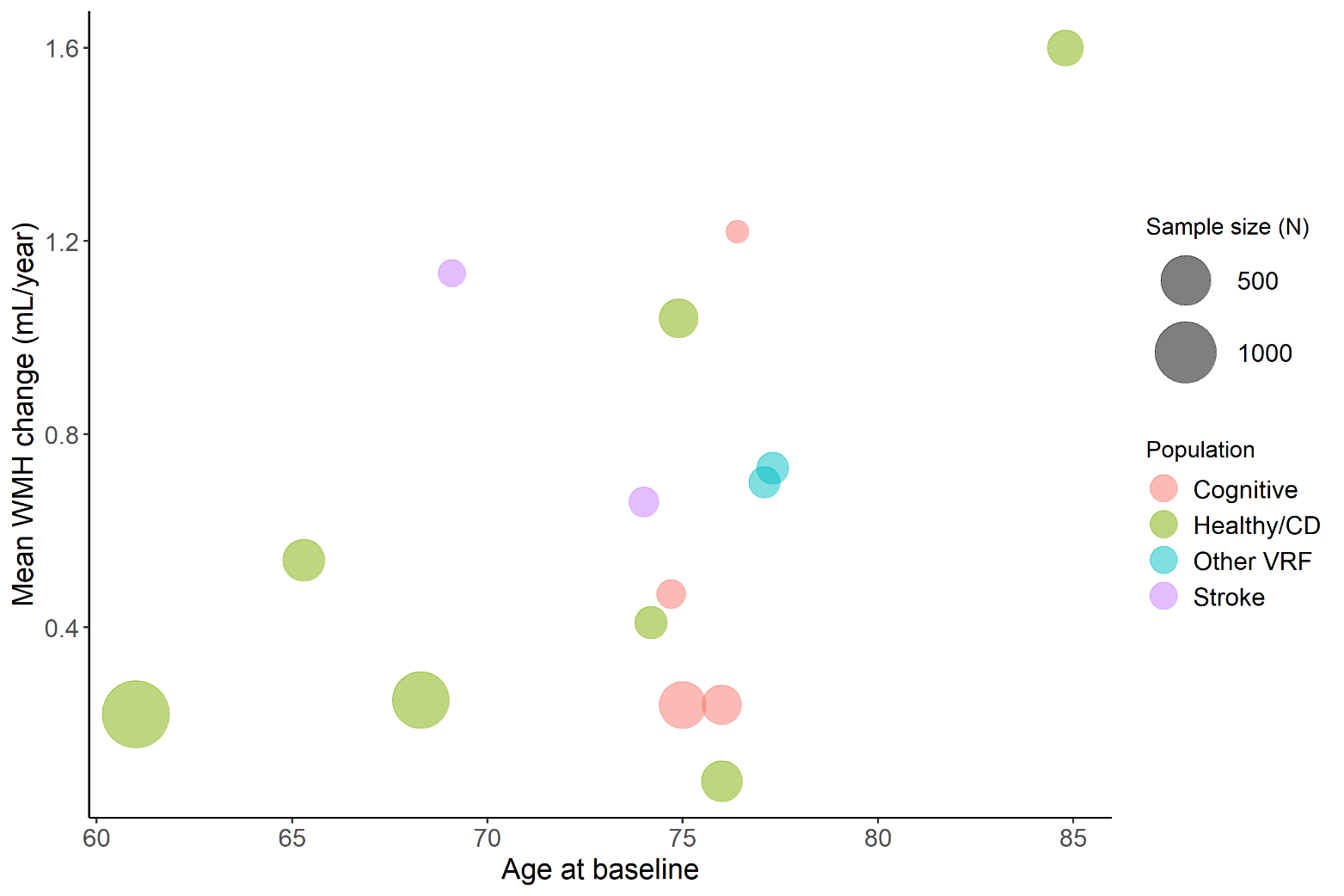
